# Mixed methods approach to examining the implementation experience of a phone-based health research survey investigating risk factors for SARS-CoV-2 infection in California

**DOI:** 10.1101/2023.02.27.23286454

**Authors:** Nozomi Fukui, Sophia S. Li, Jennifer DeGuzman, Jennifer F. Myers, John Openshaw, Anjali Sharma, James Watt, Joseph A. Lewnard, Seema Jain, Kristin L. Andrejko, Jake M. Pry, the California COVID-19 Case-Control Study Team

## Abstract

**Objective:** To describe the implementation of a test-negative design case-control study in California during the Coronavirus Disease 2019 (COVID-19) pandemic.

**Methods:** Between February 24, 2021 - February 24, 2022, 34 interviewers called 38,470 SARS-CoV-2-tested Californians to enroll 1,885 cases and 1,871 controls in a 20-minute telephone survey. We estimated adjusted odds ratios for answering the phone and consenting to participate using mixed effects logistic regression. We used a web-based anonymous survey to compile interviewer experiences.

**Results:** Cases had 1.29-fold (95% CI: 1.24–1.35) higher adjusted odds of answering the phone and 1.69-fold (1.56–1.83) higher adjusted odds of consenting to participate compared to controls. Calls placed from 4pm to 6pm had the highest adjusted odds of being answered. Interviewers who faced participants with dire need for social services or harassment experienced poor mental health.

**Conclusions:** We suggest calling during afternoons and allocating more effort towards enrolling controls when designing a case-control study. Remaining adaptive to the dynamic needs of the team is critical to a successful study, especially in a pandemic setting.

## INTRODUCTION

The Coronavirus Disease 2019 (COVID-19) pandemic called for a rapid mobilization of public health research to inform public health policy.^1^ Observational studies have played critical roles in defining COVID-19 epidemiology by identifying risk factors for infection and estimating the effectiveness of vaccines and other mitigation strategies.^2–7^ Many observational studies conducted during the COVID-19 pandemic utilized remote technologies, such as phone surveys, to safely enroll participants, however, unknown willingness to participate on this platform may pose unique challenges.^8–12^ Understanding participation patterns in phone surveys conducted during the pandemic may help optimize the implementation of future epidemiologic studies by informing sampling design strategies.

Prior to the COVID-19 pandemic, determinants of participation in phone surveys varied by disease, age, and time of day.^11,13–15^ Individuals who have or individuals adjacent to person(s) who have a history of disease are more likely to participate than individuals who are unaffected.^11^ Younger people may be more willing to answer an unknown caller, but less willing to participate in a survey with a public health official that involves disclosing sensitive information such as their recent contacts.^14^ Additionally, the time of day that individuals are called may also influence participation in the survey.^15^ The polarization of public health throughout the pandemic, including increasingly negative attitudes towards contact tracing, may limit willingness to participate in phone-based COVID-19 research.^16–19^ In the novel, dynamic context of the COVID-19 pandemic, identification of predictors of participation in observational studies using remote technologies are limited. Additionally, public health professionals report experiencing harassment, substantial mental health burdens, and burnout during the pandemic.^20^ Details regarding the toll of emotionally demanding research on researchers, especially novice researchers, is scant.^21–23^

We describe the implementation of a phone-based, test-negative design SARS-CoV-2 case-control study in California during the COVID-19 pandemic. We estimate predictors of answering the phone, subsequently enrolling in the study, and identify reasons for declining survey participation. Furthermore, we provide qualitative description of interviewer experiences to identify successes and gaps in staff support systems. These components are critical to successful and ethical study implementation and can inform the design of future epidemiologic studies conducted throughout the COVID-19 pandemic or similar pandemic settings.

## METHODS

### Study design and enrollment process

We reviewed data collected from February 24, 2021 to February 24, 2022 by the California Department of Public Health (CDPH) test-negative design case-control study that evaluated risk factors for SARS-CoV-2 infection (**Figure S1**).^6,7^ Potential case and control participants were defined as individuals with a positive and negative laboratory-confirmed SARS-CoV-2 test result, respectively. Cases and controls were individually matched by age group, birth sex, multi-county region, and SARS-CoV-2 test result window (≤7-day difference). Throughout the study period, trained interviewers used soft-phone technology with a California area code to call a list of individuals and facilitate a 20-minute survey in English or Spanish (**Figure S2**). A guiding script accompanied the semi-structured electronic survey instrument to standardize the experience of study participants (**Item S1**). Before procuring consent, interviewers alerted potential participants of the estimated 20 minutes to complete the survey.

Individuals were eligible to participate if they could provide informed consent and reported no clinical diagnosis of COVID-19 or positive test result for SARS-CoV-2 infection prior to their most recent test result. From January 6, 2022, as at-home test use increased, those with a previous (within 2 days) positive result using an at-home test became eligible. If not capable of answering questions, enrollment proceeded if a proxy respondent was available, and the potential participant gave informed consent both to participation and to have the proxy respondent answer on their behalf. For minors (<18 years old), the parent/guardian provided informed consent.

Interviewers were encouraged to enroll a case, followed by 30 or more calls to matched controls, in a repeated case-control pair format. If unsuccessful in completing a survey with a matched control within their shift, interviewers posted a request in an instant messaging platform for other interviewers to attempt enrollment in subsequent shifts. To limit recall bias, cases were excluded in the primary analysis if not matched by the 7th day. Interviewers documented the outcome (no answer, no consent, partial survey, completed survey) of each call and noted reasons for refusing participation or ending the call early.

The State of California, Health and Human Services Agency, Committee for the Protection of Human Subjects (Project Number: 2021-034) approved the study protocol.

### Implementation infrastructure

Interviewers collected data daily (excluding holidays) for an average of 10 hours per week. Research associates, promoted from interviewer roles, helped fulfill administrative and research support tasks such as maintaining databases, managing interviewer training, assigning call lists, facilitating weekly meetings, monitoring enrollment, and cultivating community.

A communication platform provided live support to interviewers who encountered questions during surveys and served as an option for private communication, group communication, and team-sourced support. Supervisors monitored the platform daily to ensure that all questions received timely responses. The online messaging platform allowed for consolidation of high volume and frequency of communication and positioned the team to easily deliver critical updates, solicit feedback on survey implementation, and detect any issues quickly.

The team met weekly to discuss study enrollment progress, check in on wellbeing, highlight interviewer accomplishments, and announce protocol or survey updates. Supervisors offered professional development opportunities during these meetings such as presentations from various public health professionals and workshops covering relevant skills and topics.

Interviewers intermittently encountered difficult conversations with participants. Team-wide, small-group, and 1-on-1 discussions about wellbeing recurred throughout the year to debrief difficult experiences, and mental wellness resources, including counseling and general support conferences across CDPH COVID-19 response sections, were advertised and encouraged.

### Interviewer recruitment and training

Interviewers were recruited from undergraduate and graduate institutions with pay (**Table S1**). Successful candidates demonstrated strong empathy, patience, good communication, interest in public health or a related field, and had prior customer service, data collection, or healthcare experience. Interviewers were required to complete a rigorous training program to ensure that they were well prepared for challenging interviews and collecting high-quality data (**Figure 1**). Due to high interviewer turnover in the first three months of the study, multiple hiring sessions occurred. With successive rounds of interviewer on-boarding we implemented a train-the-trainer approach, empowering experienced interviewers to mentor newer interviewers and respond to questions in communal message channels.

**FIGURE 1.**
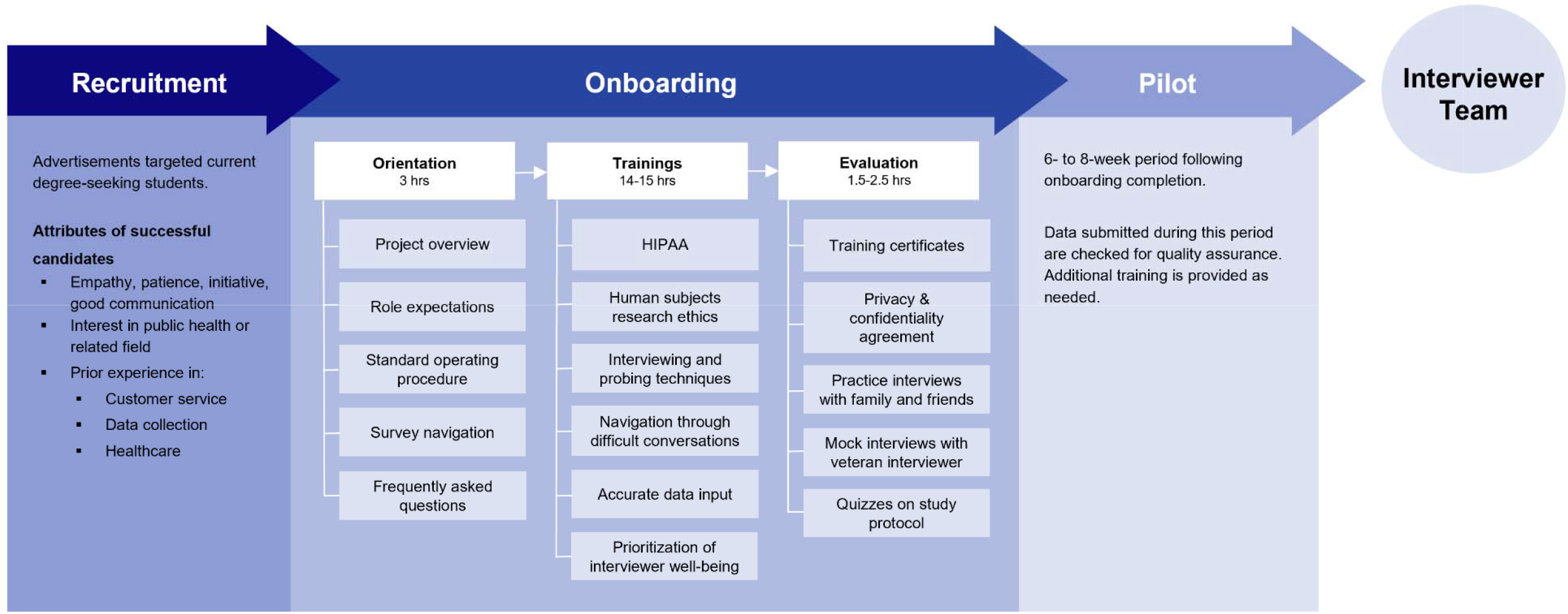
Process diagram for recruitment, onboarding, and training interviewers

### Quantitative methods

We define three cohorts representing different call outcomes: 1) individuals who answered the phone, 2) eligible individuals who consented, and 3) eligible individuals who refused to participate in the study. To estimate determinants of participation, we estimated the adjusted odds ratio of answering the phone, consenting to participate, and citing time as a reason for not participating using mixed effects logistic regression. Models included age group, birth sex, region, SARS-CoV-2 infection status, month, time of day and time of week contacted as fixed effects and allowed random effects at the interviewer level. Additionally, we assessed interaction effects between predictors by SARS-CoV-2 infection status and between time of day and time of week. Model fitting was performed to compare fit with and without inclusion of interaction terms using the Bayesian Information Criterion (**Table S2**).

All analyses were conducted with R software (version 4.1.3; R Foundation for Statistical computing). We used lme4 packages for analyses.

### Qualitative methods

From June 29 through July 12, 2022, we used an anonymous, self-administered, web-based survey to contextualize quantitative results with interviewer experiences (**Item S2**). All interviewers involved with the study were invited to participate. We compensated active interviewers for the time spent on their responses. We also reviewed weekly meeting notes for identification of themes.

## RESULTS

During the study period, we placed 38,470 calls including 15,154 (39.4%) to cases and 23,316 (60.6%) to controls (**Table 1, Figure S3**). Among the cases and controls called, 35.5% (5,383/15,154) and 31.3% (7,289/23,316) answered the phone, respectively. Of those who answered the phone, 37.2% (2,004/5,383) and 27.3% (1,991/7,289) consented to participate. Ultimately, 1,885 cases and 1,871 controls completed the survey. Over time, survey completion declined for both cases and controls despite change in calling rate (**Figure 2**).

**Table 1.** Characteristics of SARS-CoV-2 test-seekers in California who were called, answered the phone, consented to participate, and completed the telephone survey. See Table S8 for comparison to 2020 U.S. Census Bureau American Community Survey demographics

|  |  | Tested | Called |  | Answered the phone<sup>†</sup> |  | Consented to participate |  | Completed the Survey |  |
| --- | --- | --- | --- | --- | --- | --- | --- | --- | --- | --- |
|  |  | n (%) | Case | Control | Case | Control | Case | Control | Case | Control |
|  |  | N=81,980,132 | n (%) | n (%) | n (%) | n (%) | n (%) | n (%) | n (%) | n (%) |
|  |  | Case = 76428418 (93) |  |  |  |  |  |  |  |  |
|  |  | Control = 5551714 (7) | N=15154 | N=23316 | N=5081 | N=6929 | N=2004 | N=1991 | N=1885 | N=1871 |
| Sex | Male | 37594032 (45.9) | 7410 (48.9) | 10969 (47.0) | 2496 (49.1) | 3357 (48.4) | 952 (47.5) | 954 (47.9) | 898 (47.6) | 896 (47.9) |
|  | Female | 44386100 (54.1) | 7744 (51.1) | 12347 (53.0) | 2585 (50.9) | 3572 (51.6) | 1052 (52.5) | 1037 (52.1) | 987 (52.4) | 975 (52.1) |
| Age | 0 to 4 | 2357367 (2.9) | 359 (2.4) | 498 (2.1) | 130 (2.6) | 168 (2.4) | 64 (3.2) | 56 (2.8) | 59 (3.1) | 55 (2.9) |
|  | 5 to 10 | 7186063 (8.8) | 664 (4.4) | 1062 (4.6) | 234 (4.6) | 357 (5.2) | 86 (4.3) | 109 (5.5) | 84 (4.5) | 98 (5.2) |
|  | 11 to 13 | 3890275 (4.7) | 406 (2.7) | 595 (2.6) | 134 (2.6) | 181 (2.6) | 57 (2.8) | 50 (2.5) | 54 (2.9) | 47 (2.5) |
|  | 14 to 17 | 5976574 (7.3) | 772 (5.1) | 1106 (4.7) | 250 (4.9) | 332 (4.8) | 92 (4.6) | 96 (4.8) | 86 (4.6) | 87 (4.6) |
|  | 18 to 22 | 6799813 (8.3) | 1143 (7.5) | 2431 (10.4) | 413 (8.1) | 767 (11.1) | 195 (9.7) | 200 (10.0) | 176 (9.3) | 185 (9.9) |
|  | 23 to 29 | 9942434 (12.1) | 2567 (16.9) | 4260 (18.3) | 904 (17.8) | 1317 (19.0) | 383 (19.1) | 377 (18.9) | 369 (19.6) | 356 (19.0) |
|  | 30 to 39 | 13426157 (16.4) | 2830 (18.7) | 4381 (18.8) | 991 (19.5) | 1377 (19.9) | 376 (18.8) | 388 (19.5) | 358 (19.0) | 362 (19.3) |
|  | 40 to 49 | 10816874 (13.2) | 2230 (14.7) | 3396 (14.6) | 755 (14.9) | 996 (14.4) | 299 (14.9) | 288 (14.5) | 276 (14.6) | 277 (14.8) |
|  | 50 to 59 | 9699282 (11.8) | 1798 (11.9) | 2513 (10.8) | 589 (11.6) | 636 (9.2) | 211 (10.5) | 176 (8.8) | 200 (10.6) | 168 (9.0) |
|  | 60+ | 11885293 (14.5) | 2385 (15.7) | 3074 (13.2) | 681 (13.4) | 798 (11.5) | 241 (12.0) | 251 (12.6) | 223 (11.8) | 236 (12.6) |
| Region | San Francisco Bay Area | 18078093 (22.1) | 1432 (9.4) | 2317 (9.9) | 539 (10.6) | 758 (10.9) | 231 (11.5) | 228 (11.5) | 218 (11.6) | 212 (11.3) |
|  | Central Coast | 626715 (0.8) | 1642 (10.8) | 2880 (12.4) | 557 (11.0) | 742 (10.7) | 242 (12.1) | 225 (11.3) | 223 (11.8) | 218 (11.7) |
|  | Greater Sacramento Area | 2686924 (3.3) | 1617 (10.7) | 2618 (11.2) | 553 (10.9) | 867 (12.5) | 236 (11.8) | 241 (12.1) | 221 (11.7) | 225 (12.0) |
|  | Northern Sacramento Valley | 1762248 (2.1) | 1442 (9.5) | 2179 (9.3) | 472 (9.3) | 712 (10.3) | 208 (10.4) | 208 (10.4) | 197 (10.5) | 199 (10.6) |
|  | San Joaquin Valley | 6707663 (8.2) | 1957 (12.9) | 3174 (13.6) | 664 (13.1) | 917 (13.2) | 239 (11.9) | 232 (11.7) | 220 (11.7) | 219 (11.7) |
|  | Northwestern California | 840819 (1.0) | 1540 (10.2) | 2203 (9.4) | 493 (9.7) | 617 (8.9) | 213 (10.6) | 213 (10.7) | 201 (10.7) | 198 (10.6) |
|  | Sierras | 1465520 (1.8) | 1655 (10.9) | 2401 (10.3) | 529 (10.4) | 724 (10.4) | 198 (9.9) | 208 (10.4) | 189 (10.0) | 190 (10.2) |
|  | San Diego and southern border | 6692230 (8.2) | 1673 (11.0) | 2617 (11.2) | 591 (11.6) | 747 (10.8) | 214 (10.7) | 219 (11.0) | 203 (10.8) | 206 (11.0) |
|  | Greater Los Angeles Area | 43119920 (52.6) | 2196 (14.5) | 2927 (12.6) | 683 (13.4) | 845 (12.2) | 223 (11.1) | 217 (10.9) | 213 (11.3) | 204 (10.9) |
| Month | February (2021) | 4937436 (6.0) | 177 (1.2) | 228 (1.0) | 66 (1.3) | 78 (1.1) | 35 (1.7) | 30 (1.5) | 34 (1.8) | 29 (1.5) |
|  | March | 4675127 (5.7) | 1694 (11.2) | 2443 (10.5) | 651 (12.8) | 844 (12.2) | 301 (15.0) | 298 (15.0) | 272 (14.4) | 279 (14.9) |
|  | April | 4708939 (5.7) | 2009 (13.3) | 2661 (11.4) | 760 (15.0) | 869 (12.5) | 324 (16.2) | 309 (15.5) | 302 (16.0) | 289 (15.4) |
|  | May | 4051343 (4.9) | 1539 (10.2) | 2328 (10.0) | 569 (11.2) | 754 (10.9) | 224 (11.2) | 225 (11.3) | 209 (11.1) | 206 (11.0) |
|  | June | 3150746 (3.8) | 1346 (8.9) | 1683 (7.2) | 428 (8.4) | 516 (7.4) | 158 (7.9) | 156 (7.8) | 149 (7.9) | 146 (7.8) |
|  | July | 3474680 (4.2) | 1040 (6.9) | 1487 (6.4) | 350 (6.9) | 450 (6.5) | 146 (7.3) | 138 (6.9) | 137 (7.3) | 133 (7.1) |
|  | August | 6679276 (8.1) | 744 (4.9) | 1207 (5.2) | 271 (5.3) | 389 (5.6) | 116 (5.8) | 118 (5.9) | 111 (5.9) | 110 (5.9) |
|  | September | 7596972 (9.3) | 600 (4.0) | 987 (4.2) | 205 (4.0) | 324 (4.7) | 87 (4.3) | 92 (4.6) | 82 (4.4) | 85 (4.5) |
|  | October | 6647015 (8.1) | 924 (6.1) | 1884 (8.1) | 319 (6.3) | 563 (8.1) | 110 (5.5) | 119 (6.0) | 108 (5.7) | 110 (5.9) |
|  | November | 5748797 (7.0) | 1098 (7.2) | 1662 (7.1) | 345 (6.8) | 457 (6.6) | 106 (5.3) | 98 (4.9) | 100 (5.3) | 97 (5.2) |
|  | December | 7875324 (9.6) | 1344 (8.9) | 2816 (12.1) | 394 (7.8) | 682 (9.8) | 154 (7.7) | 161 (8.1) | 146 (7.7) | 149 (8.0) |
|  | January (2022) | 14487871 (18) | 1665 (11.0) | 2488 (10.7) | 467 (9.2) | 666 (9.6) | 171 (8.5) | 171 (8.6) | 166 (8.8) | 168 (9.0) |
|  | February | 7946606 (10) | 974 (6.4) | 1442 (6.2) | 256 (5.0) | 337 (4.9) | 72 (3.6) | 76 (3.8) | 69 (3.7) | 70 (3.7) |
| Time of week | Weekday |  | 12384 (81.7) | 19115 (82.0) | 4138 (81.4) | 5725 (82.6) | 1656 (82.6) | 1641 (82.4) | 1561 (82.8) | 1539 (82.3) |
|  | Weekend |  | 2770 (18.3) | 4201 (18.0) | 943 (18.6) | 1204 (17.4) | 348 (17.4) | 350 (17.6) | 324 (17.2) | 332 (17.7) |
| Time of day | 8-11am |  | 4272 (28.2) | 4833 (20.7) | 1355 (26.7) | 1336 (19.3) | 494 (24.7) | 347 (17.4) | 455 (24.1) | 331 (17.7) |
|  | 12-3pm |  | 7090 (46.8) | 11792 (50.6) | 2428 (47.8) | 3475 (50.2) | 996 (49.7) | 989 (49.7) | 945 (50.1) | 924 (49.4) |
|  | 4-6pm |  | 3483 (23.0) | 5798 (24.9) | 1198 (23.6) | 1855 (26.8) | 465 (23.2) | 563 (28.3) | 441 (23.4) | 532 (28.4) |
|  | After 6pm |  | 309 (2.0) | 893 (3.8) | 100 (2.0) | 263 (3.8) | 49 (2.4) | 92 (4.6) | 44 (2.3) | 84 (4.5) |

<sup>†</sup>This is restricted to the subpopulation of individuals who answered the phone and were eligible for the study. The 662 potential participants (360 controls and 302 cases) who answered the telephone call but were ineligible for the study and therefore excluded from this count (**Figure 1**).

**FIGURE 2.**
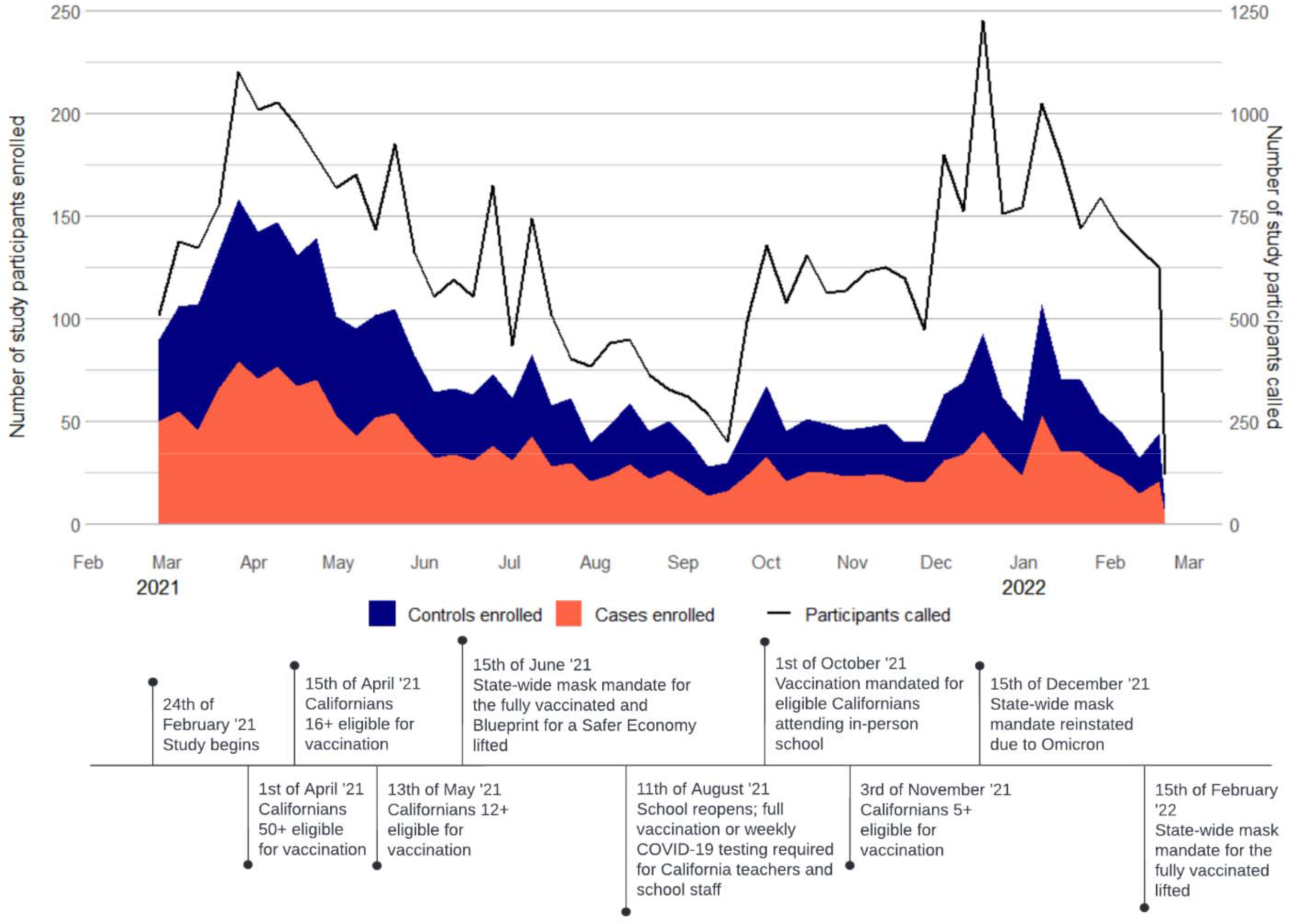
Study timeline mapped against weekly enrollment trends by case-control status

On average, interviewers placed 8 calls per case and 13 calls per control to complete a survey. Parents or guardians of children aged 0 to 4 years required fewer calls to complete a survey compared to participants in other age categories (7 calls per case and 10 calls per control) (**Figure S4**). The greatest number of calls to complete a survey for a potential participant occurred between 8am to 11am (10 calls per case and 15 calls per control). On average, the weekly calls per completed survey for a control increased over time while remaining relatively steady for cases (**Figure S5**).

During the study, there were three hiring rounds and a total of 34 interviewers. Interviewers were, on average, active for 23 weeks. 17.6% (6/34) remained active for 34-52 weeks. 32.4% (11/34) of interviewers responded to the anonymous qualitative experience survey.

### Predictors of answering the phone and consenting to participate

We found SARS-CoV-2 infection status, age, region, time of day called, and time of week called were significantly associated with answering the phone. Cases were more likely (aOR: 1.29 [95% CI: 1.24–1.35]) to answer the phone than controls (**Figure 3**). The likelihood of answering the phone was lowest among older individuals. Individuals from Central Coast (aOR: 0.77 [95% CI: 0.70, 0.85]), San Joaquin Valley (0.84 [0.77, 0.92]), and Northwestern California (0.84 [0.76, 0.93]) had the lowest adjusted odds of answering when compared to individuals in the Bay Area (**Table S3**). Calls placed after 6pm (aOR: 0.79 [0.68, 0.90]) and between 8 to 11am (0.84 [0.79–0.90]) were associated with the lowest adjusted odds of answering the phone when compared to calls placed between 4 to 6pm.

**FIGURE 3.**
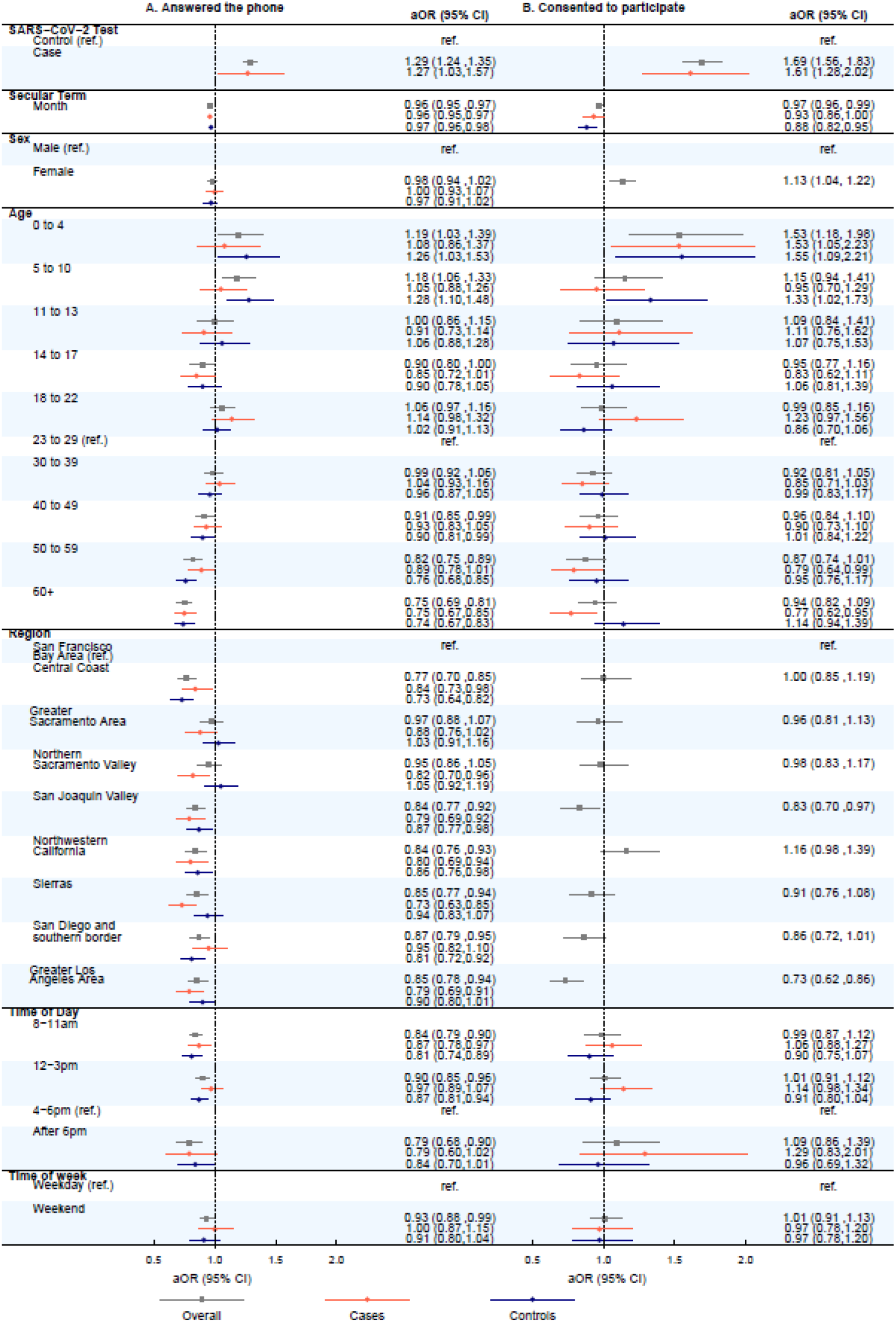
Predictors of participants answering the telephone and consenting to participate in the California COVID-19 Case Control study. We did not observe significant interaction between SARS-CoV-2 infection status and the adjusted odds of consenting to participate by region or sex. Estimates for cases and controls are not pictured for these two predictors of consenting to participate.

We also evaluated predictors of consenting to participate and found significant associations with SARS-CoV-2 infection status, age group, birth sex, and region. Cases had 1.69-fold higher adjusted odds of consenting compared to controls ([95%CI: 1.56–1.83], **Figure 3**). Women had 1.13-fold (1.04–1.22) higher adjusted odds of consenting than men. Parents or guardians of minors aged 0 to 4 were 1.53-times (1.18–1.98) more likely to consent than those aged 23 to 29. When compared with individuals from the Bay Area, individuals from San Joaquin Valley (aOR: 0.83 [0.70–0.97]) and Greater Los Angeles Area (0.73 [0.62–0.86]) were significantly less likely to consent.

Some motivations for participants to consent, as cited among interviewer reflections, were desire to contribute to public health research, relieve boredom, and express perspectives about the pandemic:

> *Participants genuinely believe their answers will help end the pandemic in some way*.
>
> *I think for the people who were inclined to not participate because of beliefs, some changed their minds when it was reframed as us wanting to make sure everyone is represented, that their voice matters, and this is a chance for them to be heard*.

We identified differences in the likelihood of answering the phone within SARS-CoV-2 infection status strata among regions (aOR for cases 0.73 [95% CI: 0.63,0.85] versus aOR for controls 0.94 [95% CI: 0.83,1.07] in the Sierras) (**Table S4**). Differences in likelihood of consenting to participate occurred within SARS-CoV-2 infection status strata among age groups (aOR for cases 0.77 [95% CI: 0.62, 0.95] versus aOR for controls 1.14 [95% CI: 0.94,1.39] aged 60 and older).

### Reasons for refusing to participate

We identified differences in the reasons for refusing to participate among 8,015 eligible individuals who answered the phone. The majority (90.9%; 7285/8015) cited insufficient time as the reason for refusing participation, with the proportion citing this reason increasing over time (**Figure S6**). Others cited language barriers (2.6%; 206/8015), lack of interest (2.3%; 184/8015), call fatigue (0.45%; 36/8015), and/or being unwell or grieving (0.45%; 36/8015) as refusal reasons (**Table S5**).

We assessed determinants of indicating insufficient time as a reason for refusing study participation. Cases were associated with a 0.44-fold (95% CI: 0.37–0.53) lower adjusted odds of citing insufficient time compared to controls (**Table S6**). Individuals aged 23 to 29 were most likely to cite insufficient time compared to all other age categories. We did not find evidence of significant associations between the time of day or the time of week the individual was called and citing time as a reason for declining participation.

Although interviewers observed that individuals often refused based on timing, they identified additional reasons, including personal beliefs, distrust, illness, and stress:

> *Some people that were politically against the public health response often declined to get interviewed or were outright confrontational*.
>
> *It is stressful for COVID-19 positive and negative cases to follow through with the interview due to sickness, worry, or even suspicions of the intents and validity of our study*.

### Sample diversity

Participants completing the survey were similar to the SARS-CoV-2 test seeking population in California across birth sex and in age groups 0-4, 18-22, 40-49, and 60+ (**Table 1, Table S7**). By design, participants were enrolled equally across each study region. The composition of study participants was roughly proportional to the state by household income and race/ethnicity (**Figure S7**).

Pandemic sentiments and behaviors, as self-reported by participants, were diverse and changed over time. Agreement with social distancing and face mask recommendations generally remained constant throughout the study period (**Figure S8**), however, anxiety about the pandemic fluctuated between 67.7% in February 2021 (44/65 participants) and 29.6% in July 2021 (84/284 participants). Participants reporting visiting two or more public indoor settings within the two weeks prior to getting tested increased from 58.5% (38/65) in February 2021 to 85.1% (126/148) by February 2022. Attendance to each type of indoor setting remained constant - except for a decrease in grocery store visits and increase in school visits (**Figure S9**). The proportion of individuals ineligible for enrollment due to previously being infected with SARS-CoV-2 increased throughout the study period (**Figure S10**).

Emotional states among participants, as encountered by interviewers, were also variable. The range of pandemic-related emotions that participants expressed included resilience, weariness, loneliness, and anger:

> *A lot of people stated that they were so tired of living through a pandemic*.
>
> *Some people angrily shared experiences about being knowingly exposed to COVID-19 by their bosses or clients during work*.

Some interviewers reported occurrences of previously vaccine-opposed participants expressing willingness to seek COVID-19 vaccination after testing positive.

> *A participant who was firmly against believing in covid ended up changing his mind after testing positive. He told me that he would get the vaccine when he could*.

### Impact on interviewer wellbeing

Of the 11 interviewers who responded to the interviewer experience survey, 63.6% (7) stated they occasionally encountered scenarios where they were compelled to search for or connect participants to social services and 18.3% (2) stated they encountered this need often (**Figure S11**). Resources pertaining to COVID-19 healthcare access, general healthcare access, housing security, food security, and financial relief were the most frequently requested. Most interviewers reported encountering grief or anger occasionally (81.8%, 9/11). 18.2% (2) reported encountering anger often.

Interviewers expressed instances of poor mental health and lingering feelings after difficult calls. Interviews where participants discussed socioeconomic and demographic burdens, pandemic hardship, grief, suffering, inequitable conditions, or acted with hostility and bullying left a particularly strong impact on interviewers. Interviewers described feelings of stress, especially when participants compelled them to fulfill social service or counselor roles.

> *Calls that I had where people shared their anxiety, confusion, fear, anger, and sadness fed into my own anxiety and negative feelings*.

Interviewers also described many encounters which instilled a sense of purpose, pride, spurred personal growth, cultivated a sense of community, expanded empathy, and uplifted moods. Notably, encounters when participants expressed appreciation, gratitude, humor, or warmth despite hardships had resounding effects on interviewers.

> *A lot of people said, “thank you for what you’re doing*.*”*… *That made me proud to be part of such an important research group*.

Interviewers also mentioned how the study provided remote career growth and employment during a time of scarce opportunities.

### Structural successes and adaptations

Feedback was frequently solicited to identify improvement opportunities. When mental health concerns surfaced, quick action was taken to strengthen structural support, community engagement, and resources. Research associates, with experience as interviewers, developed and led a robust training process that emphasized mental wellbeing and methods to navigate difficult conversations. They also compiled information on frequently requested social services and expanded on the standard operating procedure with scenario-specific protocols and responses to demands beyond interviewer duties. Active efforts to sustain a work environment that felt safe, supportive, and caring were made to better protect the mental health of interviewers.

> *We were allowed to give feedback (and our feedback was valuable and used in survey changes), and supervisors cared about our mental health over collecting data*.

Interviewers reported certain structural components as being particularly beneficial: self-assigned scheduling of shift times and weekly meetings. Self-assigned shifts allowed interviewers affected by difficult conversations to take breaks. Meeting weekly helped boost team morale, relieve isolation, and created bonding between team members:

> *The team was very supportive when I was sharing my experience, which helped show me that it is normal to feel the impact the participants may have on us whether they are at their highest or their lowest*.
>
> *Seeing the data I had helped collect be used in real time to help improve understandings of COVID inspired me to keep calling people, even when I would reach voicemail after voicemail*.
>
> *Because it was a remote job, I sometimes felt as though I was working alone, but weekly meetings helped provide that sense of teamwork*.

## DISCUSSION

Over a one-year period, during the COVID-19 pandemic, 9.8% of 38,470 individuals invited to our COVID-19 phone-based questionnaire consented to participate. Because the study was conducted across an evolving landscape of COVID-19 epidemiology and public health recommendations, flexibility to adapt protocols, exclusion criteria, and survey questions so that they remained meaningful was necessary. Results were consistent with prior research demonstrating that individuals who have a history of disease are more willing to participate in a health study than those naïve to the disease.^11^ The overall likelihood of an individual answering the phone decreased with age. Older individuals may experience more severe health burdens or reside in institutions unreachable by direct calls.^24,18^ The time of day that a potential participant was called influenced the likelihood of answering the phone, but not of consenting to participate. Results confirmed literature reporting that morning calls yield lower enrollment, indicating that strategically timing calls is crucial in maximizing enrollment efficiency.^15^

This study was successful in representing the population seeking SARS-CoV-2 testing in California, with a recruitment effort of almost 40,000 calls and a well-powered size of nearly 4,000 participants within the first year. The quality of the data allowed for identification of reasons for unsuccessful enrollment and determinants of participation. The infrastructure of the study, particularly the weekly meetings, detailed standard operating procedure documentation, and use of a messaging platform enabled quick identification of obstacles and adaptations in implementation.

Enrollment—especially of controls—became more difficult throughout the study. This difficulty may be explained by the increase in previously positive individuals but also by potentially diminished interest or perceived risk regarding the pandemic. Insufficient time was the predominant reason for refusing participation. We recommend shortening survey length or offering call-backs.

Interviewers highlighted key themes unique to the experience of remote phone-based data collection during the COVID-19 pandemic. Notably, instances of strong participant emotion and harassment especially burdened the mental health of student researchers. We believe this unforeseen consequence is novel in remote, phone-based quantitative health research and may be unique to the national context of polarized attitudes towards the COVID-19 pandemic.^25^ Proactively adapting to emerging obstacles was critical to the success of the study. Designing trainings that simulated various realistic scenarios and specific, detailed protocols for difficult encounters resulted in considerable improvement for subsequent interviewer cohorts. We suggest implementation of frequent proactive mental health check-ins, continual collection of anonymous feedback, and an exit survey for interviewers.

There are several limitations to this analysis. First, due to data constraints, we were unable to examine how socioeconomic status, race, education, occupation, and setting, such as housing, may influence the likelihood of answering the phone and consenting to participate. Second, results may not be generalizable to the broader California population, as individuals who did not seek laboratory-confirmed SARS-CoV-2 testing are excluded by design. Third, severely ill SARS-CoV-2 positive individuals, unwell individuals with comorbidities, those without stable phone service, and those cautious about phone solicitations might not be well represented in our study.

## CONCLUSIONS

We recommend placing calls during the afternoon and evening, allocating more efforts towards enrolling controls, and restricting survey length if possible. It is also imperative to consistently monitor unsuccessful recruitment efforts to detect unforeseen barriers to data collection and to ensure timely adjustments to the study protocol. Actively evaluating study implementation is vital to adapting data collection strategies when conducting a health study in a dynamic context such as a pandemic.

In summary, these findings allow researchers to strategize recruitment for future phone-based observational studies conducted amidst an evolving pandemic. We hope that our results can improve the design and implementation of future phone-based observational research.

## Data Availability

All data produced in the present study are available upon reasonable request to the authors.

## ACKNOWLEDGEMENTS

We would like to thank all participants that dedicated time to complete our survey, making the findings stemming from this research possible.

## DISCLAIMER

The findings and conclusions in this article are those of the author(s) and do not necessarily represent the views or opinions of the California Department of Public Health or the California Health and Human Services Agency.

## CONTRIBUTIONS

NF: Formal Analysis, Writing – Original Draft Preparation, Project Administration, Methodology, Visualization, Data Curation, Conceptualization

SSL: Writing – Original Draft Preparation, Visualization, Validation, Methodology

JD: Visualization, Writing – Original Draft Preparation, Conceptualization, Methodology

JM: Methodology, Conceptualization, Writing – Review & Editing

JO: Methodology, Conceptualization, Writing – Review & Editing

JW: Methodology, Conceptualization, Writing – Review & Editing, Funding Acquisition

JAL: Supervision, Methodology, Conceptualization, Writing – Review & Editing, Funding Acquisition

SJ: Methodology, Conceptualization, Writing – Review & Editing, Funding Acquisition

KLA: Supervision, Project Administration, Methodology, Writing – Review & Editing, Data Curation, Conceptualization

JMP: Supervision, Project Administration, Methodology, Writing – Review & Editing, Data Curation, Conceptualization, Funding Acquisition

AS: Writing – Review & Editing, Methodology

## FUNDING

Centers for Disease Control and Prevention, Epidemiology and Laboratory Capacity (ELC) grant 0187.0150.

## Notes

### Competing Interest Statement

The authors have declared no competing interest.

### Funding Statement

This study was funded by the Centers for Disease Control and Prevention, Epidemiology and Laboratory Capacity (ELC) building grant (number 5-NU50CK000539).

### Author Declarations

The study protocol was approved by the State of California, Health and Human Services Agency, Committee for the Protection of Human Subjects (Project Number: 2021-034).

