## Supplemental tables and figures for "Mixed methods approach to examining the implementation experience of a phone-based health research survey investigating risk factors for SARS-CoV-2 infection in California"

^§^ Members of the California COVID-19 Case-Control Study Team include: Adrian Cornejo, Amanda Lam, Amanda Moe, Amandeep Kaur, Anna Fang, Ashly Dyke, Camilla Barbaduomo, Christine Wan, Diana Nicole Morales Felipe, Diana Poindexter, Erin Xavier, Hyemin Park, Helia Samani, Jessica Ni, Julia Cheunkarndee, Mahsa Javadi, Maya Spencer, Michelle Spinosa, Miriam Bermejo, Monique Miller, Najla Dabbagh, Natalie Dassian, Nikolina Walas, Paulina Frost, Savannah Corredor, Shrey Saretha, Timothy Ho, Vivian Tran, Yang Zhou, Yasmine Abdulrahim, Zheng Dong

**Table of Contents**

| Table S1 | Characteristics of interviewers……………………………..................................................................………….. | 2 |
| --- | --- | --- |
| Table S2 | Model comparison using Bayesian Information Criterion.................................................................………….. | 3 |
| Table S3 | Predictors of answering the telephone and consenting to participate in a case-control study using mixed effects logistic regression...........................................................................................………….………….…… | 4 |
| Table S4 | Predictors of answering the telephone, consenting to participate in a case-control study, and ultimately enrolling in the study, interacted with SARS-CoV-2 test result, using mixed effects logistic regression……... | 5 |
| Table S5 | Reasons for refusal to participate in the case-control study............................................................................. | 6 |
| Table S6 | Predictors of citing time as a reason for refusing participation using mixed effects logistic regression............ | 7 |
| Table S7 | Characteristics of California population and composition of SARS-CoV-2 test seekers in California throughout the study period……………………………….................................................................…………… | 8 |
| Table S8 | 2020 U.S. Census Bureau American Community Survey Demographics of the State of California…………... | 9 |
| Figure S1 | Study regions...................................................................................................................................…………... | 10 |
| Figure S2 | Sampling process………………………………………………………………………………………….…………… | 11 |
| Figure S3 | Enrollment of participants in the California COVID-19 Case-Control Study……………………………………… | 12 |
| Figure S4 | Calls to successfully enroll a case (SARS-CoV-2 positive) or control (SARS-CoV-2 negative)……………….. | 13 |
| Figure S5 | Calls to enroll one case or control over the study period, overlayed with total cases reported across California each week……………………………………………………………………………………………………. | 14 |
| Figure S6 | Predominant reasons cited for not consenting to participate in the study, over the course of the study……… | 15 |
| Figure S7 | Comparison of household income and race/ethnicity demographics between the study population and California population...………………………………………………………………………………………………….. | 16 |
| Figure S8 | Agreement with social distancing, face mask use, general anxiety about COVID-19, and attendance at indoor public settings month of study among participants who completed the survey………………………….. | 17 |
| Figure S9 | Proportion of participants reporting attendance at indoor settings over the study period………………………. | 18 |
| Figure S10 | Area graph of population who was excluded due to previous SARS-CoV-2 positive over time………………... | 19 |
| Figure S11 | Interviewer encounters with grief, anger, and demand for social service resources……………….……………. | 20 |
| Item S1 | Survey questions and guide……………………………………………………………………………...……………. | 21 |
| Item S2 | Interviewer experience survey …………………………………………………………………………..……………. | 62 |

**Table S1.** Characteristics of interviewers

|  | **Total** | **Average num. of calls per week** | | **Average num. of complete surveys per week** | |
| --- | --- | --- | --- | --- | --- |
|  |  | < 65 calls | ≥65 calls | < 5 surveys | ≥5 surveys |
|  | *n* (%) | *n* (%) | *n* (%) | *n* (%) | *n* (%) |
|  | *N*=33 | *N*=24 | *N*=9 | *N*=15 | *N*=18 |
| Degree program |  |  |  |  |  |
| Bachelor's | 18 (54.5) | 13 (54.2) | 5 (55.6) | 9 (60.0) | 9 (50.0) |
| Master's | 13 (39.4) | 9 (37.5) | 4 (44.4) | 6 (40.0) | 7 (38.9) |
| Doctoral | 2 (6.1) | 2 (8.3) | 0 (0) | 0 (0) | 2 (11.1) |
| Customer service or outreach experience |  |  |  |  |  |
| Yes | 16 (48.5) | 11 (45.8) | 5 (55.6) | 7 (46.7) | 9 (50.0) |
| No | 17 (51.5) | 13 (54.2) | 4 (44.4) | 8 (53.3) | 9 (50.0) |
| Research or data collection experience |  |  |  |  |  |
| Yes | 23 (69.7) | 16 (66.7) | 7 (77.8) | 9 (60.0) | 14 (77.8) |
| No | 10 (30.3) | 8 (33.3) | 2 (22.2) | 6 (40.0) | 4 (22.2) |
| Healthcare or patient-facing experience |  |  |  |  |  |
| Yes | 19 (57.6) | 12 (50.0) | 7 (77.8) | 7 (46.7) | 12 (66.7) |
| No | 14 (42.4) | 12 (50.0) | 2 (22.2) | 8 (53.3) | 6 (33.3) |

**Table S2. Model Comparison Using BIC**

| **Model** | **Outcome** | | | |
| --- | --- | --- | --- | --- |
|  | Answering | Consenting | Completing survey | Citing lack of time as a reason for not consenting |
|  | BIC | BIC | BIC | BIC |
| Logistic regression, no interactions | 48140 | 15161 | 14862 | 4194 |
| Random effects model, no interactions | 47949 | 14990 | 14352 | 4041 |
| Logistic regression, interaction with case status | 48349 | 15290 | 14993 | 4279 |
| Random effects model, interaction with case status | 48158 | 15110 | 14323 | 4126 |

**Table S3.** Predictors of answering the telephone and consenting to participate in a case-control study using mixed effects logistic regression. Model comparisons using BIC are listed in **Table S2**.

|  |  | **Answer the telephone** | | **Consented to participate** | | **Completed the survey** | |
| --- | --- | --- | --- | --- | --- | --- | --- |
|  |  | OR (95% CI) | aOR (95% CI) | OR (95% CI) | aOR (95% CI) | OR (95% CI) | aOR (95% CI) |
| SARS-CoV-2 Test |  |  |  |  |  |  |  |
|  | Control | ref. | ref. | ref. | ref. | ref. | ref. |
|  | Case | 1.27 (1.22, 1.33) | 1.29 (1.24, 1.35) | 1.67 (1.54, 1.81) | 1.69 (1.56, 1.83) | 1.64 (1.52, 1.78) | 1.66 (1.53 ,1.80) |
| Secular Term | Month | 0.97 (0.96, 0.98) | 0.96 (0.95, 0.97) | 0.97 (0.96, 0.99) | 0.97 (0.96, 0.99) | 0.98 (0.97, 1.00) | 0.98 (0.97 ,1.00) |
| Sex |  |  |  |  |  |  |  |
|  | Male | ref. | ref. | ref. | ref. | ref. | ref. |
|  | Female | 0.96 (0.92, 1.01) | 0.98 (0.94, 1.02) | 1.11 (1.03, 1.20) | 1.13 (1.04, 1.22) | 1.10 (1.02, 1.19) | 1.11 (1.03 ,1.21) |
| Age |  |  |  |  |  |  |  |
|  | 0 to 4 | 1.17 (1.00, 1.36) | 1.19 (1.03, 1.39) | 1.46 (1.13, 1.88) | 1.53 (1.18, 1.98) | 1.44 (1.12, 1.86) | 1.51 (1.16 ,1.96) |
|  | 5 to 10 | 1.18 (1.05, 1.32) | 1.18 (1.06, 1.33) | 1.07 (0.88, 1.30) | 1.15 (0.94, 1.41) | 1.03 (0.85, 1.26) | 1.10 (0.90 ,1.35) |
|  | 11 to 13 | 1.01 (0.87, 1.16) | 1.00 (0.86, 1.15) | 1.05 (0.81, 1.35) | 1.09 (0.84, 1.41) | 1.04 (0.80, 1.34) | 1.08 (0.83 ,1.40) |
|  | 14 to 17 | 0.91 (0.81, 1.01) | 0.90 (0.80, 1.00) | 0.96 (0.79, 1.17) | 0.95 (0.77, 1.16) | 0.92 (0.75, 1.12) | 0.90 (0.74 ,1.11) |
|  | 18 to 22 | 1.04 (0.96, 1.14) | 1.06 (0.97, 1.16) | 0.97 (0.84, 1.13) | 0.99 (0.85, 1.16) | 0.92 (0.79, 1.07) | 0.94 (0.80 ,1.09) |
|  | 23 to 29 | ref. | ref. | ref. | ref. | ref. | ref. |
|  | 30 to 39 | 0.99 (0.92, 1.06) | 0.99 (0.92, 1.06) | 0.92 (0.81, 1.04) | 0.92 (0.81, 1.05) | 0.91 (0.80, 1.03) | 0.91 (0.80 ,1.03) |
|  | 40 to 49 | 0.91 (0.84, 0.98) | 0.91 (0.85, 0.99) | 0.96 (0.84, 1.10) | 0.96 (0.84, 1.10) | 0.94 (0.82, 1.08) | 0.94 (0.82 ,1.08) |
|  | 50 to 59 | 0.81 (0.75, 0.88) | 0.82 (0.75, 0.89) | 0.90 (0.77, 1.05) | 0.87 (0.74, 1.01) | 0.90 (0.77, 1.05) | 0.87 (0.74 ,1.01) |
|  | 60+ | 0.75 (0.69, 0.81) | 0.75 (0.69, 0.81) | 0.97 (0.84, 1.12) | 0.94 (0.82, 1.09) | 0.94 (0.82, 1.09) | 0.92 (0.79 ,1.06) |
| Region |  |  |  |  |  |  |  |
|  | San Francisco Bay Area | ref. | ref. | ref. | ref. | ref. | ref. |
|  | Central Coast | 0.75 (0.68, 0.82) | 0.77 (0.70, 0.85) | 1.01 (0.86, 1.19) | 1.00 (0.85, 1.19) | 1.01 (0.86, 1.20) | 1.01 (0.85 ,1.20) |
|  | Greater Sacramento Area | 0.97 (0.88, 1.06) | 0.97 (0.88, 1.07) | 0.95 (0.80, 1.11) | 0.96 (0.81, 1.13) | 0.94 (0.80, 1.11) | 0.96 (0.81 ,1.13) |
|  | Northern Sacramento Valley | 0.93 (0.85, 1.03) | 0.95 (0.86, 1.05) | 0.98 (0.83, 1.17) | 0.98 (0.83, 1.17) | 1.00 (0.84, 1.19) | 1.00 (0.84 ,1.20) |
|  | San Joaquin Valley | 0.83 (0.76, 0.91) | 0.84 (0.77, 0.92) | 0.83 (0.71, 0.98) | 0.83 (0.70, 0.97) | 0.83 (0.71, 0.98) | 0.83 (0.70 ,0.98) |
|  | Northwestern California | 0.79 (0.72, 0.87) | 0.84 (0.76, 0.93) | 1.14 (0.96, 1.36) | 1.16 (0.98, 1.39) | 1.14 (0.96, 1.36) | 1.16 (0.97 ,1.39) |
|  | Sierras | 0.82 (0.74, 0.90) | 0.85 (0.77, 0.94) | 0.91 (0.77, 1.08) | 0.91 (0.76, 1.08) | 0.91 (0.77, 1.08) | 0.92 (0.77 ,1.09) |
|  | San Diego and southern border | 0.86 (0.79, 0.95) | 0.87 (0.79, 0.95) | 0.88 (0.74, 1.03) | 0.86 (0.72, 1.01) | 0.89 (0.75, 1.05) | 0.87 (0.73 ,1.03) |
|  | Greater Los Angeles Area | 0.84 (0.77, 0.92) | 0.85 (0.78, 0.94) | 0.74 (0.63, 0.87) | 0.73 (0.62, 0.86) | 0.76 (0.64, 0.89) | 0.75 (0.63 ,0.88) |
| Time of Day |  |  |  |  |  |  |  |
|  | 8-11am | 0.87 (0.82, 0.94) | 0.84 (0.79, 0.90) | 1.06 (0.94, 1.20) | 0.99 (0.87, 1.12) | 1.02 (0.90, 1.15) | 0.95 (0.84 ,1.08) |
|  | 12-3pm | 0.92 (0.87, 0.97) | 0.90 (0.85, 0.96) | 1.03 (0.93, 1.14) | 1.01 (0.91, 1.12) | 1.01 (0.91, 1.12) | 0.99 (0.90 ,1.10) |
|  | 4-6pm | ref. | ref. | ref. | ref. | ref. | ref. |
|  | After 6pm | 0.77 (0.67, 0.89) | 0.79 (0.68, 0.90) | 1.03 (0.81, 1.31) | 1.09 (0.86, 1.39) | 0.97 (0.76, 1.23) | 1.02 (0.80 ,1.31) |
| Time of week |  |  |  |  |  |  |  |
|  | Weekday | ref. | ref. | ref. | ref. | ref. | ref. |
|  | Weekend | 0.95 (0.90, 1.01) | 0.93 (0.88, 0.99) | 1.02 (0.91, 1.13) | 1.01 (0.91, 1.13) | 1.01 (0.91, 1.13) | 1.02 (0.91 ,1.14) |

**Table S4.** Predictors of answering the telephone, consenting to participate in a case-control study, and ultimately enrolling in the study, interacted with SARS-CoV-2 test result, using mixed effects logistic regression. Model comparisons using BIC are listed in **Table S2**.

|  |  | **Answer the telephone** | | **Consent to participate** | | **Complete the survey** | |
| --- | --- | --- | --- | --- | --- | --- | --- |
|  |  | Cases | Controls | Cases | Controls | Cases | Controls |
|  |  | *aOR (95% CI)* | *aOR (95% CI)* | *aOR (95% CI)* | *aOR (95% CI)* | *aOR (95% CI)* | *aOR (95% CI)* |
| SARS-CoV-2 Test |  |  |  |  |  |  |  |
|  | Control | ref. | ref. | ref. | ref. | ref. | ref. |
|  | Case | 1.27 (1.03,1.57) | - | 1.61 (1.28,2.02) | - | 1.69 (1.34,2.12) | - |
| Secular Term | Month | 0.96 (0.95,0.97) | 0.97 (0.96,0.98) | 0.93 (0.86,1.00) | 0.88 (0.82,0.95) | 0.96 (0.89,1.04) | 0.90 (0.84,0.97) |
| Sex |  |  |  |  |  |  |  |
|  | Male | ref. | ref. | ref. | ref. | ref. | ref. |
|  | Female | 1.00 (0.93,1.07) | 0.97 (0.91,1.02) | - | - | - | - |
| Age |  |  |  |  |  |  |  |
|  | 0 to 4 | 1.08 (0.86,1.37) | 1.26 (1.03,1.53) | 1.53 (1.05,2.23) | 1.55 (1.09,2.21) | 1.39 (0.95,2.03) | 1.64 (1.15,2.34) |
|  | 5 to 10 | 1.05 (0.88,1.26) | 1.28 (1.10,1.48) | 0.95 (0.70,1.29) | 1.33 (1.02,1.73) | 0.96 (0.71,1.30) | 1.23 (0.94,1.61) |
|  | 11 to 13 | 0.91 (0.73,1.14) | 1.06 (0.88,1.28) | 1.11 (0.76,1.62) | 1.07 (0.75,1.53) | 1.07 (0.73,1.57) | 1.08 (0.75,1.55) |
|  | 14 to 17 | 0.85 (0.72,1.01) | 0.90 (0.78,1.05) | 0.83 (0.62,1.11) | 1.06 (0.81,1.39) | 0.80 (0.59,1.08) | 1.01 (0.76,1.33) |
|  | 18 to 22 | 1.14 (0.98,1.32) | 1.02 (0.91,1.13) | 1.23 (0.97,1.56) | 0.86 (0.70,1.06) | 1.09 (0.85,1.38) | 0.84 (0.68,1.04) |
|  | 23 to 29 | ref | ref | ref | ref | ref | ref |
|  | 30 to 39 | 1.04 (0.93,1.16) | 0.96 (0.87,1.05) | 0.85 (0.71,1.03) | 0.99 (0.83,1.17) | 0.84 (0.69,1.01) | 0.97 (0.81,1.15) |
|  | 40 to 49 | 0.93 (0.83,1.05) | 0.90 (0.81,0.99) | 0.90 (0.73,1.10) | 1.01 (0.84,1.22) | 0.84 (0.68,1.02) | 1.04 (0.86,1.25) |
|  | 50 to 59 | 0.89 (0.78,1.01) | 0.76 (0.68,0.85) | 0.79 (0.64,0.99) | 0.95 (0.76,1.17) | 0.78 (0.62,0.97) | 0.96 (0.77,1.20) |
|  | 60+ | 0.75 (0.67,0.85) | 0.74 (0.67,0.83) | 0.77 (0.62,0.95) | 1.14 (0.94,1.39) | 0.73 (0.59,0.90) | 1.13 (0.93,1.39) |
| Region |  |  |  |  |  |  |  |
|  | San Francisco Bay Area | ref. | ref. | ref. | ref. | ref. | ref. |
|  | Central Coast | 0.84 (0.73,0.98) | 0.73 (0.64,0.82) | - | - | - | - |
|  | Greater Sacramento Area | 0.88 (0.76,1.02) | 1.03 (0.91,1.16) | - | - | - | - |
|  | Northern Sacramento Valley | 0.82 (0.70,0.96) | 1.05 (0.92,1.19) | - | - | - | - |
|  | San Joaquin Valley | 0.79 (0.69,0.92) | 0.87 (0.77,0.98) | - | - | - | - |
|  | Northwestern California | 0.80 (0.69,0.94) | 0.86 (0.76,0.98) | - | - | - | - |
|  | Sierras | 0.73 (0.63,0.85) | 0.94 (0.83,1.07) | - | - | - | - |
|  | San Diego and southern border | 0.95 (0.82,1.10) | 0.81 (0.72,0.92) | - | - | - | - |
|  | Greater Los Angeles Area | 0.79 (0.69,0.91) | 0.90 (0.80,1.01) | - | - | - | - |
| Time of Day |  |  |  |  |  |  |  |
|  | 8-11am | 0.87 (0.78,0.97) | 0.81 (0.74,0.89) | 1.06 (0.88,1.27) | 0.90 (0.75,1.07) | 0.99 (0.82,1.19) | 0.88 (0.74,1.06) |
|  | 12-3pm | 0.97 (0.89,1.07) | 0.87 (0.81,0.94) | 1.14 (0.98,1.34) | 0.91 (0.80,1.04) | 1.13 (0.96,1.32) | 0.89 (0.78,1.03) |
|  | 4-6pm | ref | ref | ref | ref | ref | ref |
|  | After 6pm | 0.79 (0.60,1.02) | 0.84 (0.70,1.01) | 1.29 (0.83,2.01) | 0.96 (0.69,1.32) | 1.17 (0.75,1.83) | 0.90 (0.64,1.25) |
| Time of week |  |  |  |  |  |  |  |
|  | Weekday | ref. | ref. | ref. | ref. | ref. | ref. |
|  | Weekend | 1.00 (0.87,1.15) | 0.91 (0.80,1.04) | 0.97 (0.78,1.20) | 0.97 (0.78,1.20) | 0.91 (0.70,1.17) | 1.02 (0.80,1.30) |

**Table S5.** Reasons for refusal to participate in the case-control study

|  |  | Timing or duration | Language | Not interested | Call fatigue | Unwell or grieving | Other |
| --- | --- | --- | --- | --- | --- | --- | --- |
|  | 8,015 | N= 7285 | N=206 | N=184 | N=36 | N=36 | N=268 |
| Sex | Male | 3564 (48.9) | 105 (51.0) | 96 (52.2) | 21 (58.3) | 18 (50.0) | 143 (53.4) |
|  | Female | 3721 (51.1) | 101 (49.0) | 88 (47.8) | 15 (41.7) | 18 (50.0) | 125 (46.6) |
| Age |  |  |  |  |  |  |  |
|  | 0 to 4 | 163 (2.2) | 4 (1.9) | 5 (2.7) | 0 (0.0) | 0 (0.0) | 6 (2.2) |
|  | 5 to 10 | 371 (5.1) | 10 (4.9) | 1 (0.5) | 2 (5.6) | 0 (0.0) | 12 (4.5) |
|  | 11 to 13 | 180 (2.5) | 9 (4.4) | 3 (1.6) | 1 (2.8) | 2 (5.6) | 13 (4.9) |
|  | 14 to 17 | 345 (4.7) | 7 (3.4) | 14 (7.6) | 2 (5.6) | 1 (2.8) | 25 (9.3) |
|  | 18 to 22 | 736 (10.1) | 6 (2.9) | 23 (12.5) | 6 (16.7) | 0 (0.0) | 14 (5.2) |
|  | 23 to 29 | 1389 (19.1) | 10 (4.9) | 34 (18.5) | 3 (8.3) | 4 (11.1) | 21 (7.8) |
|  | 30 to 39 | 1470 (20.2) | 33 (16.0) | 38 (20.7) | 11 (30.6) | 9 (25.0) | 43 (16.0) |
|  | 40 to 49 | 1031 (14.2) | 50 (24.3) | 27 (14.7) | 4 (11.1) | 8 (22.2) | 44 (16.4) |
|  | 50 to 59 | 723 (9.9) | 44 (21.4) | 21 (11.4) | 6 (16.7) | 7 (19.4) | 37 (13.8) |
|  | 60+ | 877 (12.0) | 33 (16.0) | 18 (9.8) | 1 (2.8) | 5 (13.9) | 53 (19.8) |
| Region |  |  |  |  |  |  |  |
|  | San Francisco Bay Area | 727 (10.0) | 40 (19.4) | 15 (8.2) | 3 (8.3) | 3 (8.3) | 50 (18.7) |
|  | Central Coast | 761 (10.4) | 24 (11.7) | 17 (9.2) | 4 (11.1) | 2 (5.6) | 24 (9.0) |
|  | Greater Sacramento Area | 858 (11.8) | 23 (11.2) | 29 (15.8) | 3 (8.3) | 2 (5.6) | 28 (10.4) |
|  | Northern Sacramento Valley | 714 (9.8) | 9 (4.4) | 22 (12.0) | 3 (8.3) | 6 (16.7) | 14 (5.2) |
|  | San Joaquin Valley | 992 (13.6) | 34 (16.5) | 31 (16.8) | 9 (25.0) | 6 (16.7) | 38 (14.2) |
|  | Northwestern California | 630 (8.6) | 6 (2.9) | 17 (9.2) | 1 (2.8) | 6 (16.7) | 24 (9.0) |
|  | Sierras | 790 (10.8) | 2 (1.0) | 20 (10.9) | 8 (22.2) | 5 (13.9) | 22 (8.2) |
|  | San Diego and southern border | 813 (11.2) | 33 (16.0) | 17 (9.2) | 4 (11.1) | 2 (5.6) | 36 (13.4) |
|  | Greater Los Angeles Area | 1000 (13.7) | 35 (17.0) | 16 (8.7) | 1 (2.8) | 4 (11.1) | 32 (11.9) |
| SARS-CoV-2 Test |  |  |  |  |  |  |  |
|  | Positive (Case) | 4611 (63.3) | 77 (37.4) | 97 (52.7) | 7 (19.4) | 10 (27.8) | 136 (50.7) |
|  | Negative (Control) | 2674 (36.7) | 129 (62.6) | 87 (47.3) | 29 (80.6) | 26 (72.2) | 132 (49.3) |
| Time of week |  |  |  |  |  |  |  |
|  | Weekday | 5981 (82.1) | 169 (82.0) | 153 (83.2) | 32 (88.9) | 31 (86.1) | 200 (74.6) |
|  | Weekend | 1304 (17.9) | 37 (18.0) | 31 (16.8) | 4 (11.1) | 5 (13.9) | 68 (25.4) |
| Time of day |  |  |  |  |  |  |  |
|  | 8-11am | 1687 (23.2) | 71 (34.5) | 35 (19.0) | 5 (13.9) | 5 (13.9) | 47 (17.5) |
|  | 12-3pm | 3562 (48.9) | 95 (46.1) | 97 (52.7) | 24 (66.7) | 19 (52.8) | 121 (45.1) |
|  | 4-6pm | 1852 (25.4) | 36 (17.5) | 41 (22.3) | 7 (19.4) | 5 (13.9) | 84 (31.3) |
|  | After 6pm | 184 (2.5) | 4 (1.9) | 11 (6.0) | 0 (0.0) | 7 (19.4) | 16 (6.0) |

**Table S6.** Predictors of citing time as a reason for refusing participation using mixed effects logistic regression.

Model comparisons using BIC are listed in **Table S2**.

|  |  | **Refused due to lack of time** | |
| --- | --- | --- | --- |
|  |  | OR (95% CI) | aOR (95% CI) |
| SARS-CoV-2 Test |  |  |  |
| S12 | Control | ref. | ref. |
|  | Case | 0.45 (0.38, 0.53) | 0.44 (0.37, 0.52) |
| Secular Term | Month | 1.23 (1.19, 1.27) | 2.17 (1.90, 2.49) |
| Sex |  |  |  |
|  | Male | ref. | ref. |
|  | Female | 1.11 (0.94, 1.30) | 0.95 (0.80, 1.13) |
| Age |  |  |  |
|  | 0 to 4 | 0.64 (0.35, 1.16) | 0.61 (0.32, 1.15) |
|  | 5 to 10 | 0.74 (0.45, 1.20) | 0.73 (0.44, 1.20) |
|  | 11 to 13 | 0.34 (0.21, 0.56) | 0.28 (0.17, 0.47) |
|  | 14 to 17 | 0.40 (0.27, 0.60) | 0.41 (0.27, 0.63) |
|  | 18 to 22 | 0.80 (0.55, 1.17) | 0.74 (0.50, 1.09) |
|  | 23 to 29 | ref. | ref. |
|  | 30 to 39 | 0.53 (0.39, 0.72) | 0.54 (0.40, 0.74) |
|  | 40 to 49 | 0.35 (0.26, 0.48) | 0.34 (0.25, 0.47) |
|  | 50 to 59 | 0.32 (0.23, 0.44) | 0.31 (0.22, 0.44) |
|  | 60+ | 0.38 (0.27, 0.52) | 0.37 (0.27, 0.52) |
| Region |  |  |  |
|  | San Francisco Bay Area | ref. | ref. |
|  | Central Coast | 1.57 (1.12, 2.20) | 1.78 (1.25, 2.55) |
|  | Greater Sacramento Area | 1.70 (1.23, 2.35) | 1.57 (1.12, 2.20) |
|  | Northern Sacramento Valley | 2.03 (1.40, 2.93) | 1.96 (1.33, 2.90) |
|  | San Joaquin Valley | 1.49 (1.10, 2.01) | 1.46 (1.06, 2.01) |
|  | Northwestern California | 2.48 (1.72, 3.56) | 2.51 (1.71, 3.67) |
|  | Sierras | 3.20 (2.24, 4.57) | 4.45 (3.04, 6.50) |
|  | San Diego and southern border | 1.51 (1.11, 2.07) | 1.65 (1.19, 2.29) |
|  | Greater Los Angeles Area | 1.67 (1.22, 2.28) | 1.85 (1.33, 2.56) |
| Time of Day |  |  |  |
|  | 8-11am | 0.87 (0.68, 1.12) | 1.03 (0.78, 1.35) |
|  | 12-3pm | 0.93 (0.75, 1.14) | 0.94 (0.75, 1.18) |
|  | 4-6pm | ref. | ref. |
|  | After 6pm | 0.57 (0.37, 0.87) | 0.60 (0.38, 0.95) |
| Time of week |  |  |  |
|  | Weekday | ref. | ref. |
|  | Weekend | 0.87 (0.71, 1.07) | 1.08 (0.86, 1.36) |

**Table S7.** Characteristics of California population and composition of SARS-CoV-2 test seekers in California throughout the study period

See **Table S8** for comparison to 2020 U.S. Census Bureau American Community Survey demographics

|  |  | **Total** | **Feb ‘21** | **Mar** | **Apr** | **May** | **Jun** | **Jul** | **Aug** | **Sept** | **Oct** | **Nov** | **Dec** | **Jan ‘22** | **Feb** |
| --- | --- | --- | --- | --- | --- | --- | --- | --- | --- | --- | --- | --- | --- | --- | --- |
|  |  | *n* (%) | *n* (%) | *n* (%) | *n* (%) | *n* (%) | *n* (%) | *n* (%) | *n* (%) | *n* (%) | *n* (%) | *n* (%) | *n* (%) | *n* (%) | *n* (%) |
|  |  | *N=81980132* | *N=4937436* | *N=4675127* | *N=4708939* | *N=4051343* | *N=3150746* | *N=3474680* | *N=6679276* | *N=7596972* | *N=6647015* | *N=5748797* | *N=7875324* | *N=14487871* | *N=7946606* |
| Sex | Male | 37,594,032 (46) | 2,294,346 | 2,220,269 | 2,227,238 | 1,888,488 | 1,480,990 | 1,618,991 | 3,018,887 | 3,438,661 | 3,042,297 | 2,654,983 | 3,573,752 | 6,484,130 | 3,651,000 |
|  | Female | 44386100 (54) | 2,643,090 | 2,454,858 | 2,481,701 | 2,162,855 | 1,669,756 | 1,855,689 | 3,660,389 | 4,158,311 | 3,604,718 | 3,093,814 | 4,301,572 | 8,003,741 | 4,295,606 |
| Age | 0 to 4 | 2,357,367 (3) | 65,297 | 62,796 | 70,994 | 73,581 | 71,003 | 108,905 | 215,754 | 262,781 | 236,333 | 204,379 | 265,892 | 477,279 | 242,373 |
|  | 5 to 10 | 7,186,063 (9) | 107,164 | 140,634 | 192,545 | 210,842 | 157,839 | 180,747 | 536,077 | 857,301 | 727,498 | 592,069 | 758,081 | 1,601,310 | 1,123,956 |
|  | 11 to 13 | 3,890,275 (5) | 61,418 | 73,979 | 108,392 | 114,748 | 91,467 | 92,013 | 279,280 | 438,766 | 388,831 | 315,326 | 398,814 | 893,487 | 633,754 |
|  | 14 to 17 | 5,976,574 (7) | 116,119 | 221,369 | 353,288 | 312,579 | 147,047 | 133,728 | 445,465 | 652,511 | 540,383 | 432,812 | 571,013 | 1,253,645 | 796,615 |
|  | 18 to 22 | 6,799,813 (8) | 471,335 | 448,773 | 437,438 | 334,813 | 228,084 | 242,260 | 500,866 | 671,373 | 599,169 | 535,300 | 569,878 | 1,121,963 | 638,561 |
|  | 23 to 29 | 9,942,434 (12) | 767,344 | 687,325 | 635,944 | 524,142 | 408,583 | 491,105 | 847,990 | 862,410 | 764,040 | 661,745 | 960,796 | 1,596,227 | 734,783 |
|  | 30 to 39 | 13,426,157 (16) | 970,652 | 874,971 | 831,840 | 705,225 | 567,256 | 676,379 | 1,171,083 | 1,192,623 | 1,041,643 | 903,727 | 1,299,066 | 2,186,418 | 1,005,274 |
|  | 40 to 49 | 10,816,874 (13) | 776,952 | 709,184 | 697,804 | 584,529 | 489,525 | 520,725 | 908,546 | 923,740 | 803,907 | 701,617 | 1,016,869 | 1,834,189 | 849,287 |
|  | 50 to 59 | 9,699,282 (12) | 744,655 | 678,916 | 649,206 | 549,056 | 449,067 | 454,395 | 782,452 | 782,377 | 690,063 | 616,973 | 902,778 | 1,596,367 | 802,977 |
|  | 60+ | 11,885,293 (14) | 856,500 | 777,180 | 731,488 | 641,828 | 540,875 | 574,423 | 991,763 | 953,090 | 855,148 | 784,849 | 1,132,137 | 1,926,986 | 1,119,026 |
| Region | San Francisco Bay Area | 18,078,093 (22) | 1,213,856 | 1,240,965 | 1,209,779 | 978,831 | 731,702 | 797,742 | 1,454,278 | 1,623,178 | 1,436,045 | 1,265,857 | 1,621,636 | 2,939,122 | 1,565,102 |
|  | Central Coast | 626,715 (1) | 51,047 | 46,060 | 43,132 | 34,609 | 25,569 | 24,766 | 48,210 | 50,058 | 41,079 | 37,237 | 53,034 | 112,141 | 59,773 |
|  | Greater Sacramento Area | 2,686,924 (3) | 165,853 | 174,544 | 176,898 | 152,798 | 120,169 | 141,032 | 251,508 | 268,852 | 230,521 | 185,667 | 229,802 | 398,391 | 190,889 |
|  | Northern Sacramento Valley | 1,762,248 (2) | 90,803 | 123,124 | 112,763 | 103,620 | 72,390 | 72,521 | 139,966 | 177,508 | 173,442 | 144,364 | 155,540 | 250,158 | 146,049 |
|  | San Joaquin Valley | 6,707,663 (8) | 497,675 | 475,929 | 450,173 | 379,547 | 294,740 | 307,234 | 603,127 | 661,823 | 569,085 | 468,623 | 499,820 | 994,105 | 505,782 |
|  | Northwestern California | 840,819 (1) | 57,268 | 58,465 | 51,739 | 46,431 | 38,781 | 42,848 | 94,033 | 95,266 | 74,990 | 58,898 | 57,922 | 103,197 | 60,981 |
|  | Sierras | 1,465,520 (2) | 103,319 | 113,003 | 107,225 | 84,833 | 72,948 | 79,323 | 140,098 | 142,437 | 124,834 | 104,221 | 112,322 | 186,045 | 94,912 |
|  | San Diego and southern border | 6,692,230 (8) | 406,738 | 357,591 | 389,801 | 342,377 | 284,853 | 296,626 | 559,927 | 634,888 | 580,314 | 496,177 | 713,226 | 1,143,121 | 486,591 |
|  | Greater Los Angeles Area | 43,119,920 (53) | 2,350,877 | 2,085,446 | 2,167,429 | 1,928,297 | 1,509,594 | 1,712,588 | 3,388,129 | 3,942,962 | 3,416,705 | 2,987,753 | 4,432,022 | 8,361,591 | 4,836,527 |
| Test | Positive | 5,551,714 (7) | 225,563 | 89,858 | 69,644 | 42,095 | 33,122 | 176,619 | 405,924 | 261,530 | 177,472 | 152,572 | 602,406 | 2,868,215 | 446,694 |
|  | Negative | 76,428,418 (93) | 4,711,873 | 4,585,269 | 4,639,295 | 4,009,248 | 3,117,624 | 3,298,061 | 6,273,352 | 7,335,442 | 6,469,543 | 5,596,225 | 7,272,918 | 11,619,656 | 7,499,912 |

**Table S8.** 2020 U.S. Census Bureau American Community Survey Demographics of the State of California

|  |  | **2020 Census** |
| --- | --- | --- |
|  |  | *n* (%) |
|  |  | *N=*39,346,023 |
| Sex |  |  |
|  | Male | 19562882 (50) |
|  | Female | 19783141 (50) |
| Age |  |  |
|  | 0 to 4 | 2409082 (6) |
|  | 5 to 9 | 2431647 (6) |
|  | 10 to 14 | 2597443 (7) |
|  | 15 to 19 | 2548072 (6) |
|  | 20 to 24 | 2694636 (7) |
|  | 25 to 29 | 3084036 (8) |
|  | 30 to 39 | 5634057 (14) |
|  | 40 to 49 | 5060920 (13) |
|  | 50 to 59 | 4987445 (13) |
|  | 60+ | 7133941 (18) |
| Region |  |  |
|  | San Francisco Bay Area | 8048227 (20) |
|  | Central Coast | 1160389 (3) |
|  | Greater Sacramento Area | 1537948 (4) |
|  | Northern Sacramento Valley | 729684 (12) |
|  | San Joaquin Valley | 4225143 (11) |
|  | Northwestern California | 550503 (1) |
|  | Sierras | 932557 (2) |
|  | San Diego and southern border | 3504550 (9) |
|  | Greater Los Angeles Area | 18657022 (47) |

**Figure S1**. Study Regions

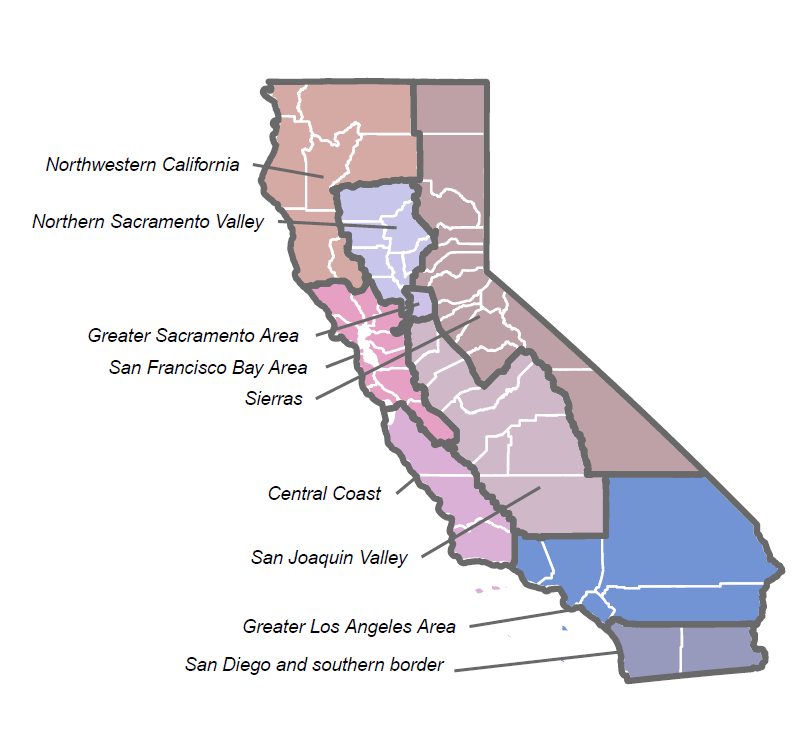

**Figure S2**. Sampling process

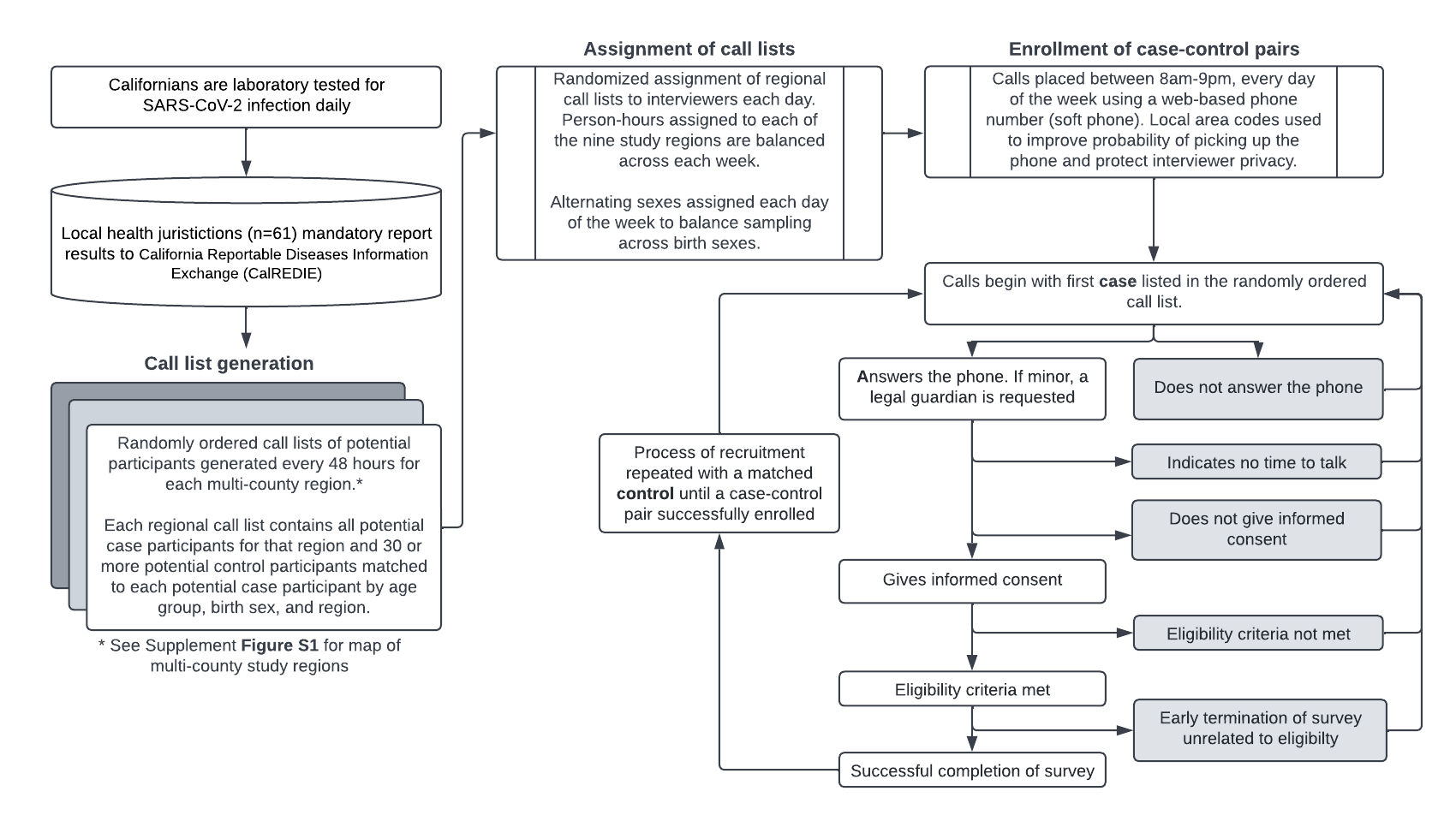

**Figure S3.** Enrollment of participants in the California COVID-19 Case-Control Study

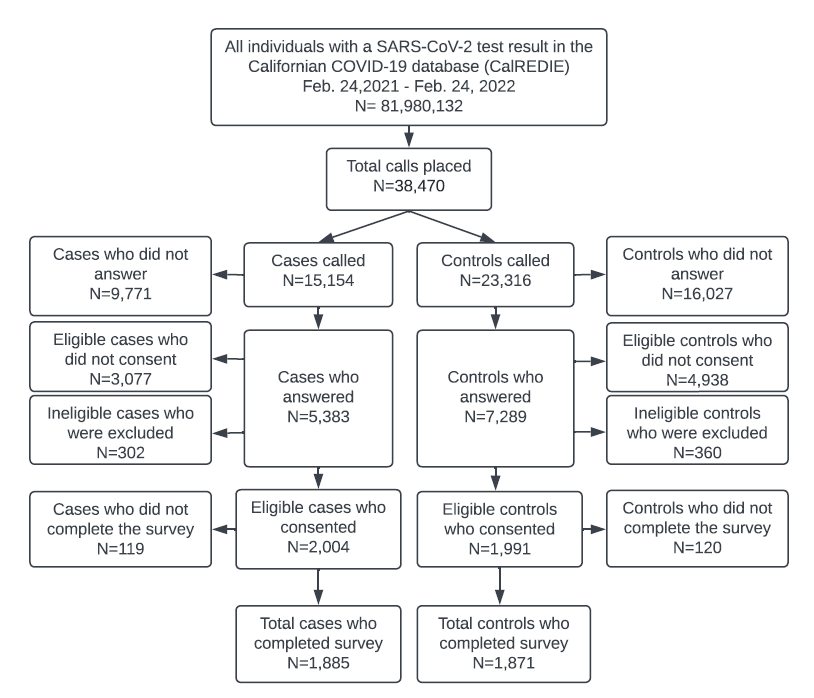

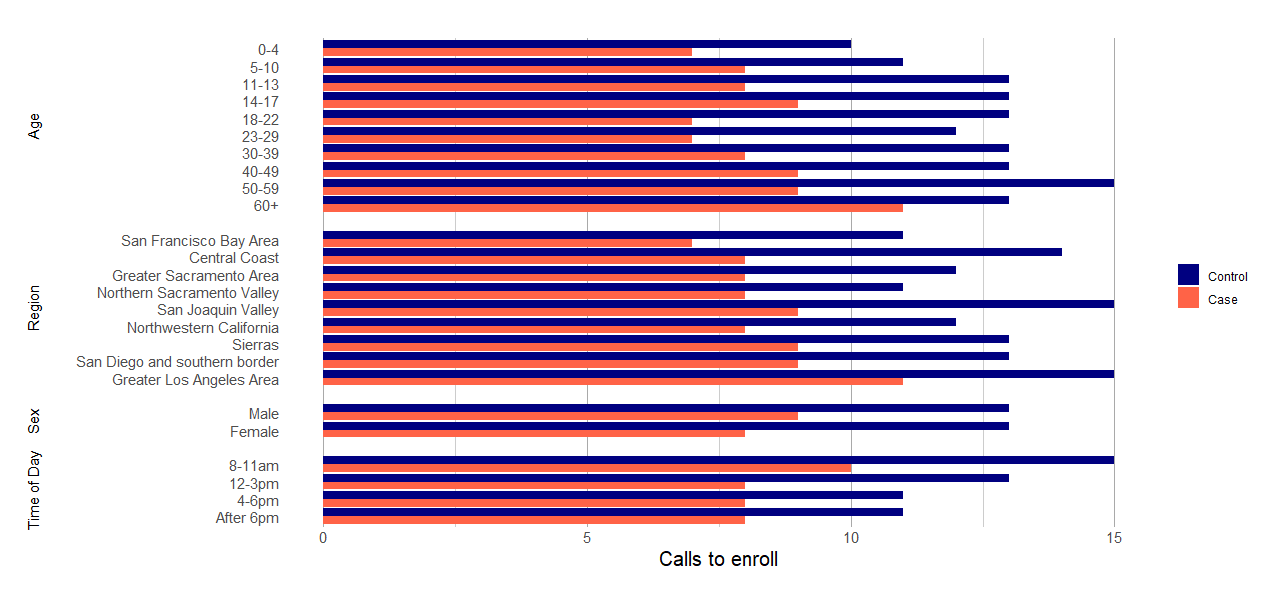
**Figure S4.** Calls to successfully enroll a case (SARS-CoV-2 positive) or control (SARS-CoV-2 negative)

**Figure S5.** Calls to enroll one case or control over the study period, overlayed with total cases reported across California each week.

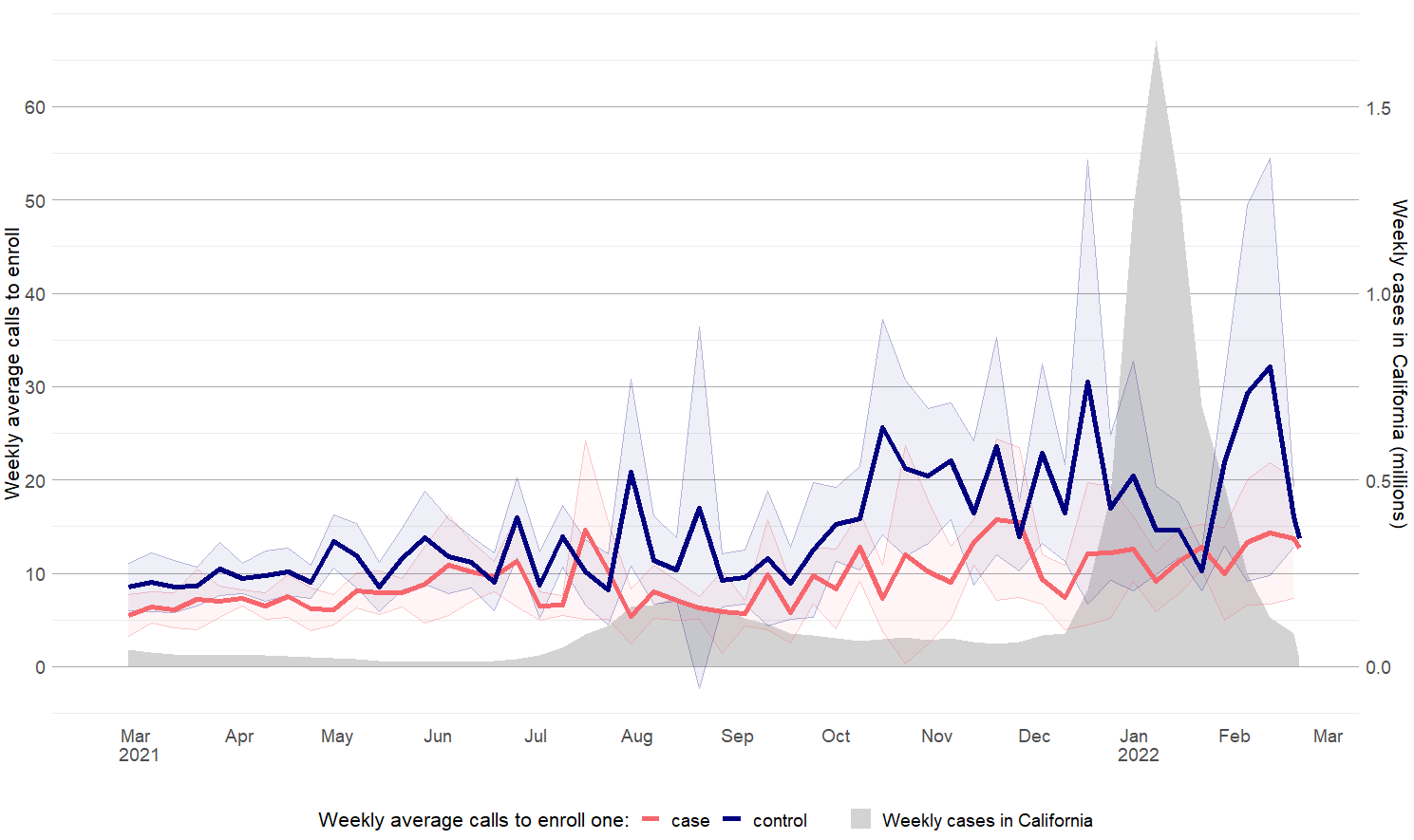

SARS-CoV-2 case data is from the California COVID-19 State Dashboard. California Department of Public Health. (2020–2022). COVID-19 Time-Series Metrics by County and State. California Open Data Portal. https://data.ca.gov/dataset/covid-19-time-series-metrics-by-county-and-state.

**Figure S6.** Predominant reasons cited for not consenting to participate in the study, over the course of the study

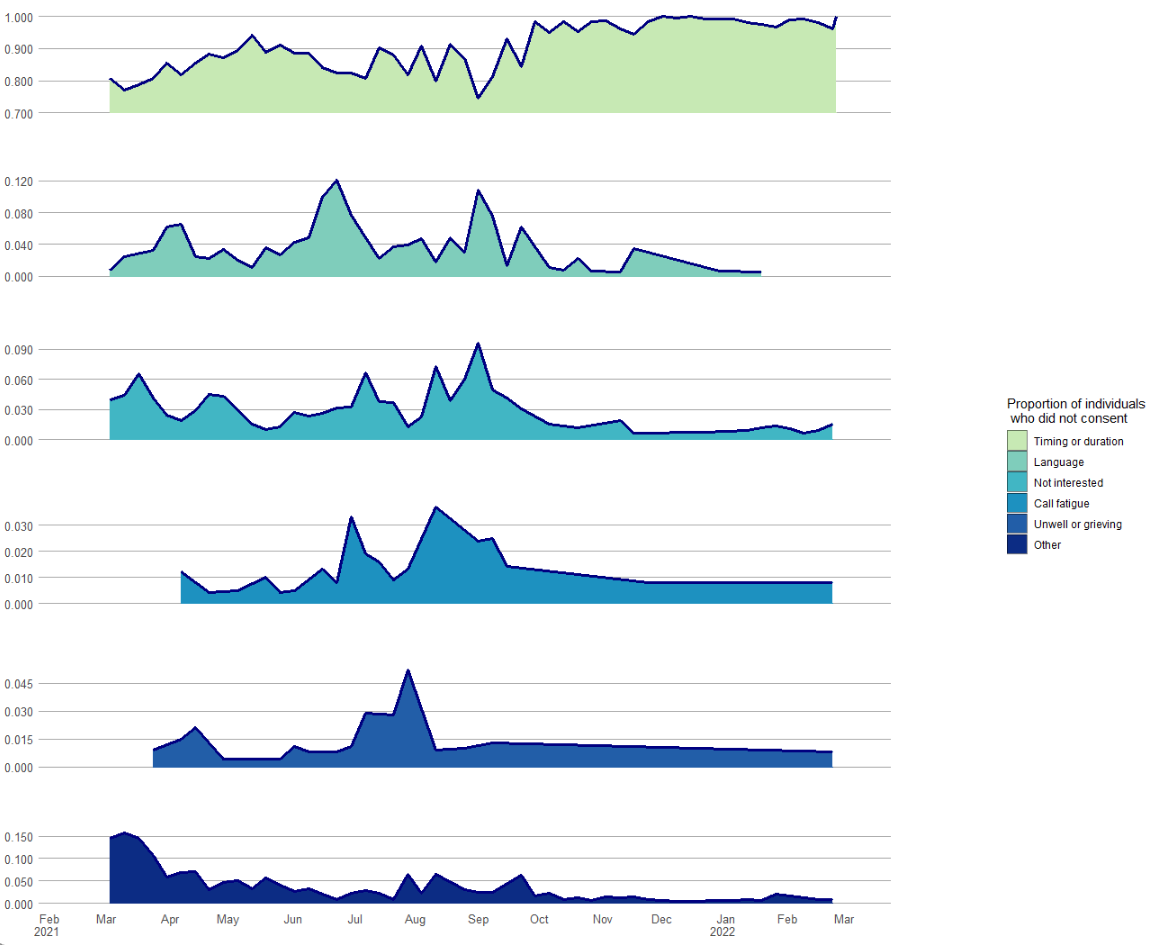

Note: This data was restricted to surveys where eligible potential participants declined informed consent.

Scale for the percent of potential participants who did not consent to participate in the study who cited each reason for declining participation differs for each plot.

**Figure S7.** Comparison of household income and race/ethnicity demographics between the study population and California population.

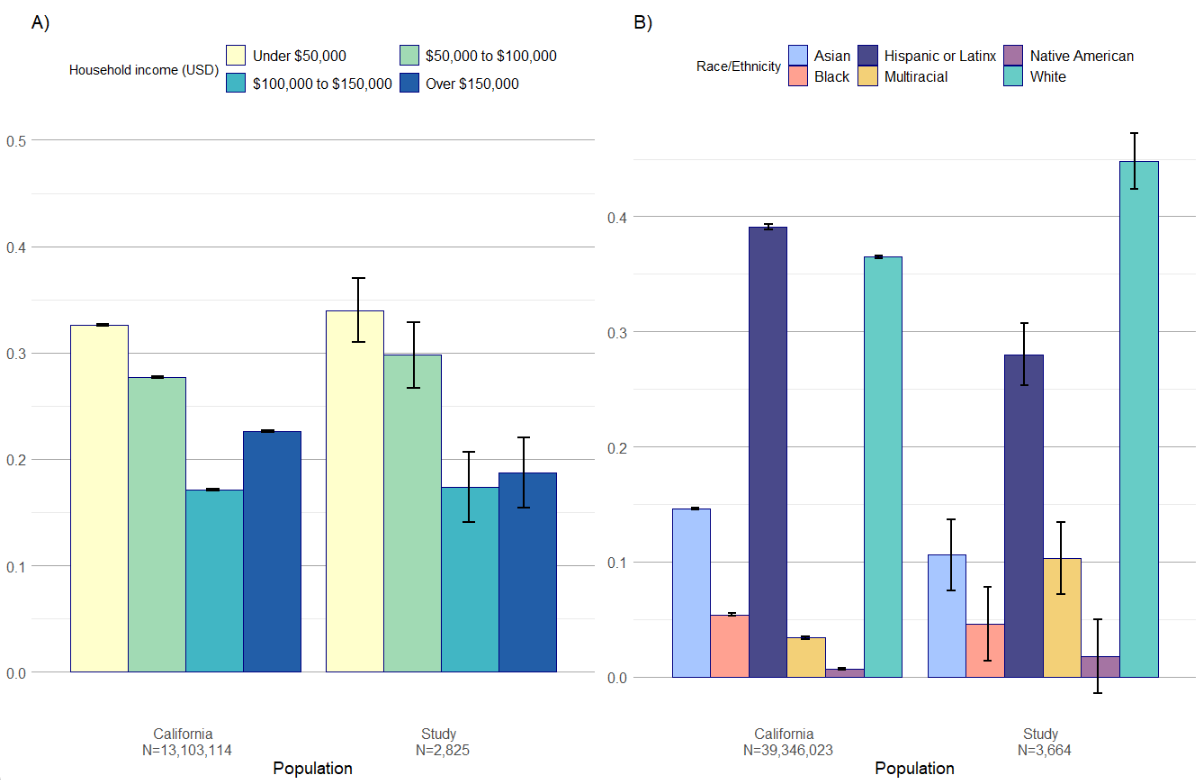

B) The California population represented is from the 2020 U.S. Census Bureau American Community Survey 5-year estimates. The study population represented includes all participants who completed the survey and provided a response to the race/ethnicity question (97.6% of 3756, or 3664 individuals). Native Hawaiians and Pacific Islanders were included in the Native American category in the study sample.

A) The California population represented is from the 2020 U.S. Census Bureau American Community Survey 5-year estimates. The study population represented includes all participants who completed the survey and provided a response to the income question (75.2% of 3756, or 2825 individuals).

**Figure S8.** Agreement with social distancing, face mask use, general anxiety about COVID-19, and attendance at indoor public settings month of study among participants who completed the survey

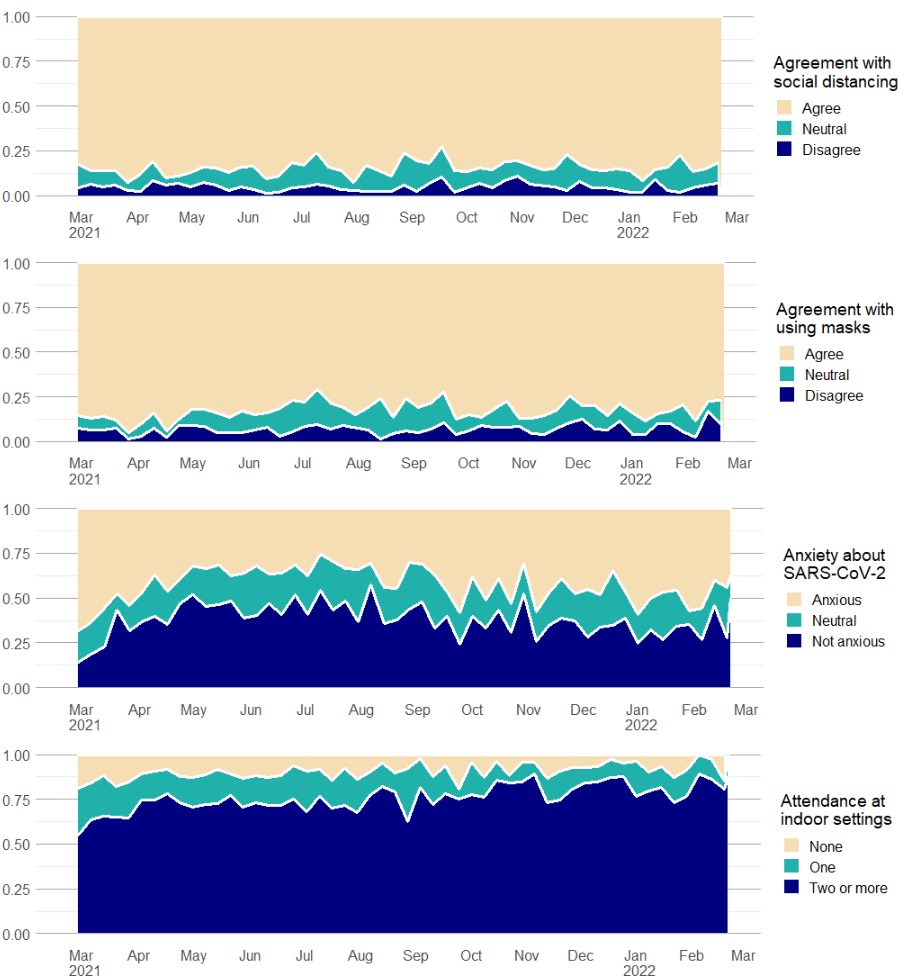

**Figure S9.** Proportion of participants reporting attendance at various indoor settings over the study period

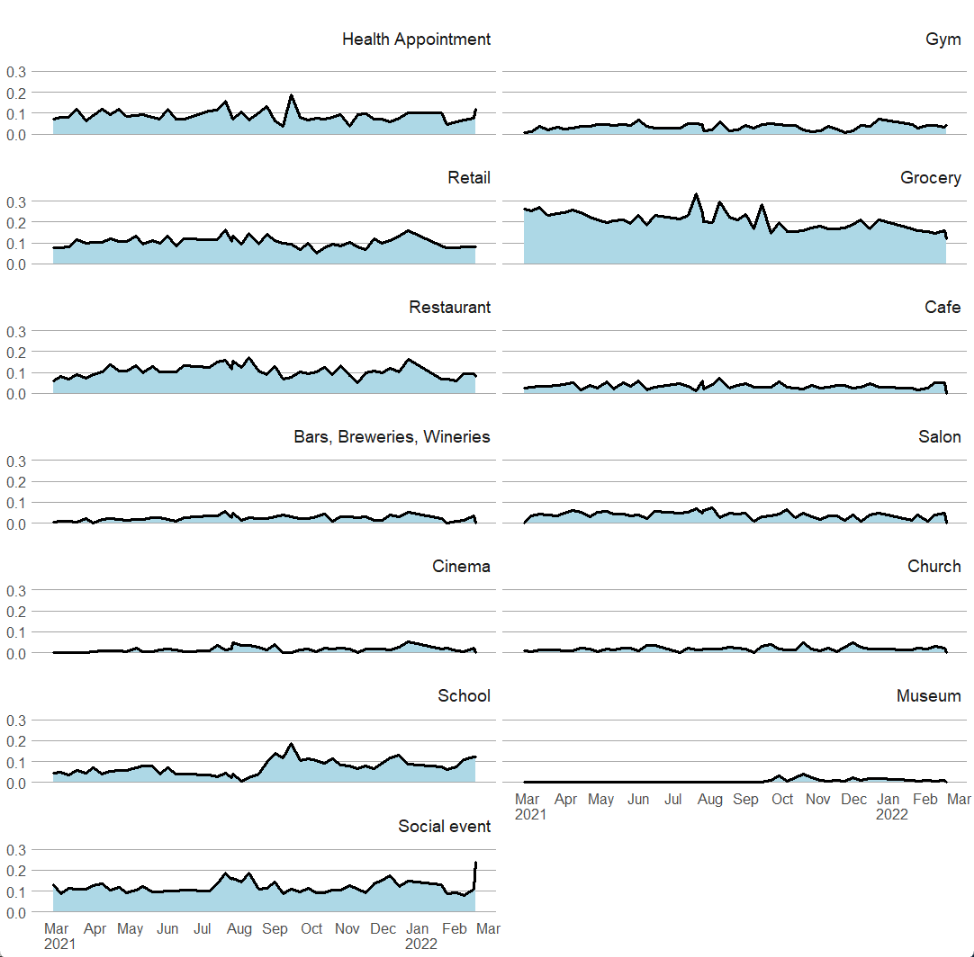

**Figure S10.** Area graph of population who was excluded due to previous SARS-CoV-2 positive over time

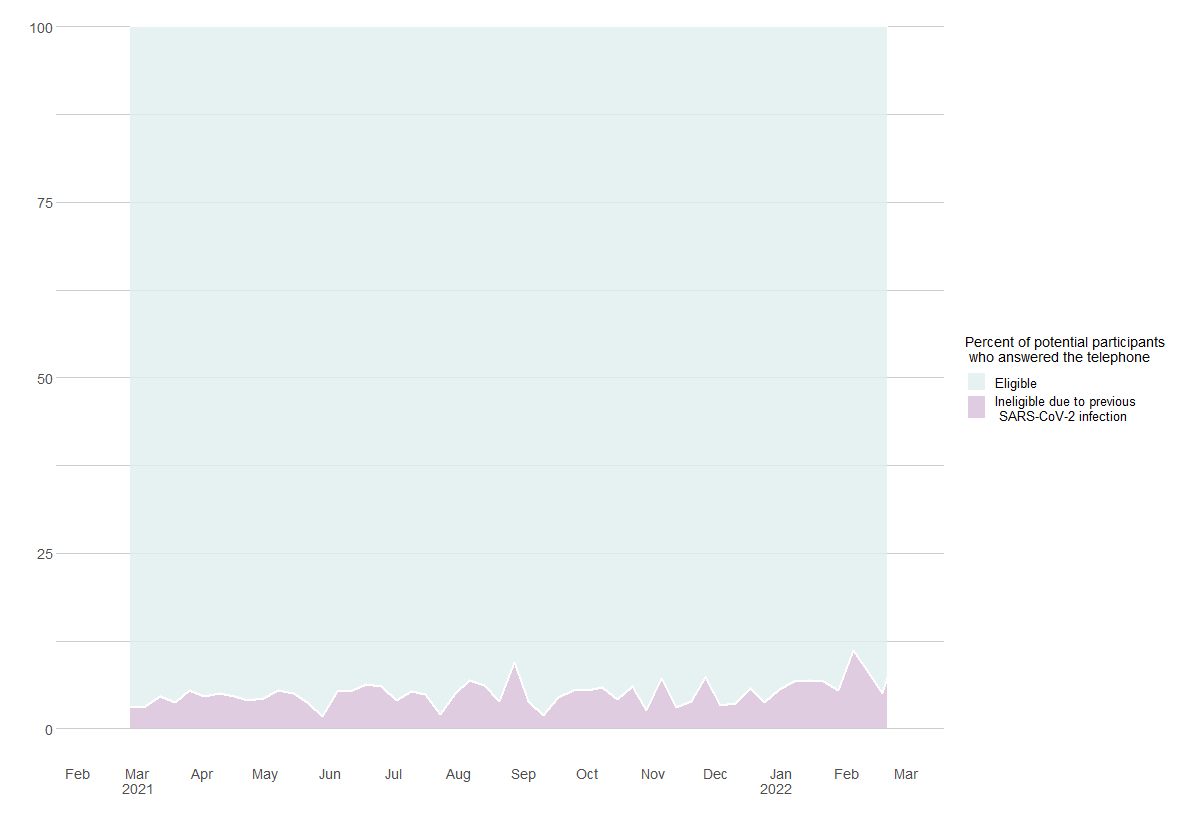

Note: Restricted to potential participants who answered the telephone.

**Figure S11.** Interviewer encounters with grief, anger, and demand for social service resources.
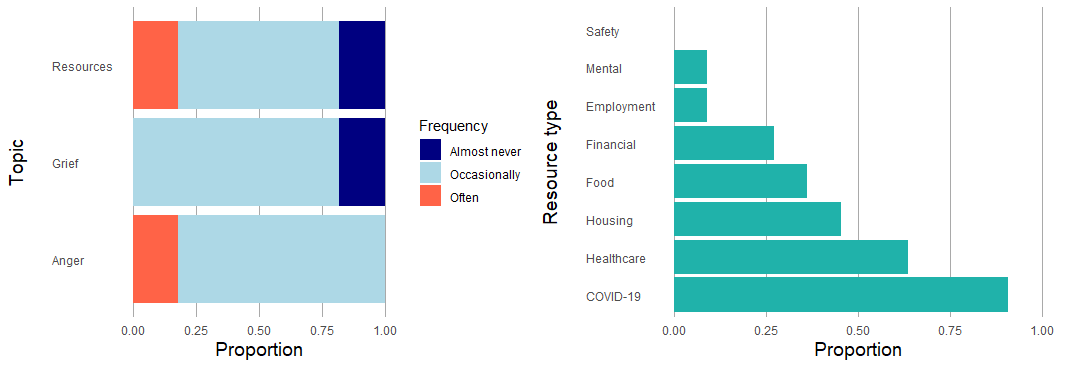

B) Note: Interviewers could select more than one resource type in their responses.

A) The topic of resources, grief, and anger are pertaining to the following three questions: How often did the people you interviewed ask you for resources or confide something concerning that compelled you to look for /share resources? How often did you encounter grief during calls? How often did you encounter anger during calls?

**Item S1.** Survey questions and guide.

COVID-19 Case-Control- Investigator Form - Pilot

Survey Flow

Standard: Information about case/ control (10 Questions)

Block: Section 1: Introduction (147 Questions)

Standard: Final block (9 Questions)

Start of Block: Information about case/ control

Please select your first and last name

▼

Please paste the case-control ID: 
NOTE: this is the "LinkLog" in column A of the spreadsheet

________________________________________________________________

Please paste the case-control ID again: 
NOTE: this is the "LinkLog" in column A of the spreadsheet

________________________________________________________________

Please select the region for the case-control:

▼ Bay Area (1) ... Southern California (9)

Please select the sex of the case-control:

- Female (1)
- Male (2)
- Not listed (3)

Are you calling a case or control?

- Case (1)
- Control (2)

Please select the age of the case-control you are interviewing

▼ 0 (1) ... 100 (101)

Please write the date that the case-control test was ADMINISTERED (MM/DD/YYYY)
This is the "specimen_collection_date" on the excel document

________________________________________________________________

Please write the date that occurred 14 days prior to the the date above (MM/DD/YYYY)

________________________________________________________________

Note to interviewer: items in italics are not intended to be communicated to the interviewee and are solely included here for the interviewers reference. Items in [blue brackets] will help you flow through the survey depending on how the interviewee responds. Sections in **BOLD** are to be communicated to the interviewer. While it is important to stay on script as much as possible to prevent bias, make sure you sound like a human and are adjusting language for the script appropriately based off of previous responses. All questions will have the option for “don’t know” or “refuse” but you should only select these if the participant explicitly states they do not know or refuse.

End of Block: Information about case/ control

Start of Block: Section 1: Introduction

*Please select "Nobody answered the phone" if the call did not go through, nobody answered the phone, the call went to voicemail, or you were unable to speak to the correct person. Otherwise, proceed with introductions*

- Nobody answered the phone (2)

**SECTION 1: INTRODUCTION (~2 min)** 
 **1.     Hello, my name is {interviewer name} and I am calling on behalf of California Department of Public Health to ask some questions regarding [NAME]’s recent COVID-19 test on {test date}** .

**2.** *Make sure you’re on the phone with the correct person*.   *If case is a child under 18y, make sure you are speaking to a parent/ guardian:*

**2a. Am I speaking to** [NAME]'s **parent or guardian?**

[If yes, proceed to **section 2**]

[If no, proceed to 2b]

**2b.** **Can you please pass the phone to** [NAME]'s parent or guardian?

[If yes, proceed to **section 2**]

[If no, end call by clicking the back button and selecting "Nobody answered the phone"]

*If case is someone older than 18y:*

**2c. Am I speaking to [NAME]?**

[If yes, proceed to **section 2**]

[If no, proceed to 2d]

**2d.** **Can you please pass the phone to [NAME]?**

[If yes, proceed to **section 2**]

[If no, end call by clicking the back button and selecting "Nobody answered the phone"]

**SECTION 2: ASSENT  (~1 min)**

If you are speaking to a parent or correct person for the first time, add your name and affiliation before starting at 1. Tailor the survey to change "Did you...." to "Did your child..."  Hello, my name is {interviewer name} and I am calling on behalf of California Department of Public Health.  1.     Hi! I am calling because I am hoping to interview you as part of a study that's trying to better understand the spread of COVID-19. This interview takes around 15 minutes. I was wondering if [YOUR or INSERT CHILD’S NAME] have time to chat?"

INTERVIEWER: pause and wait for person to confirm that they are still on the line, check YES if they say they are willing to chat

If they do not have time, select NO

- Yes I have some time to chat (1)
- No (if you select this option, it will terminate the survey!) (2)

So before we start, I want to make sure you understand that everything I ask you is confidential, protected by California’s strict privacy laws, and is only being used to inform public health. Your answers will not be shared with any other federal, state, or local authorities, and you're welcome to decline to answer any question. We anticipate this will take about 20 minutes. I know that sounds like a bit, but we really appreciate your time and your answers will help better understand COVID-19 and hopefully improve our response the pandemic. Do you understand the information I have just shared with you?

INTERVIEWER:
 check “yes” if the respondent answers yes and if you deem the respondent to be competent to proceed with consent and interviewing; check “no” and thank the respondent for their time if the respondent says no or if you deem the respondent is not competent to proceed with consent and interviewing.]

 If it seems like the person needs a proxy respondent due to not speaking well enough English or being too sick, you may ask "**Is there anyone who can help you answer my questions?".** If you get the proxy respondent on the phone, re-introduce yourself by starting at the top of Section 2 with "Hello, my name is..." and add at the end**, "Can you help answer questions on [insert name of case/control's] behalf?"**    NOTE that a proxy respondent must be over the age of 14.

- Yes I understand the information (1)
- No I do not understand this information (ex. due to language barrier and no proxy respondent available) (4)

**2.** 
  [If participant is answering on their own behalf AND they are older then 18]
           **Great, thank you! To confirm, are you willing to participate in this interview?**     [If participant is a child older than 14, answering on their own behalf, first ask for consent from the parent for the child to answer the survey]
           Great, thank you! I want to let you know that y**our child [INSERT CHILD'S NAME] may answer questions on their own behalf. Are you willing to allow [INSERT CHILD’S NAME] to participate in this** interview**?** If not, you can answer questions on their behalf. *Interviewer: if the child older than 14 joins the call, make sure to reintroduce yourself and explain the purpose of the survey.*    [If participant is a child younger than 14, and adult is answering on their behalf]
           **Great, thank you! Are you willing to answer questions about [INSERT CHILD’S NAME]'s recent exposures as part of this** interview**?** 

 [If a proxy respondent will answer on behalf of the study participant]
 If you are able, I would suggest putting the phone on speakerphone during this interview, so [insert name/ relationship of proxy respondent] can help you.            **[INSERT NAME OF CASE-CONTROL], are you willing to participate in this** interview**?**
            [INSERT NAME OF CASE-CONTROL], d**o you consent to allow [NAME OF PROXY RESPONDENT] to answer my questions during this interview. Please stay close by [NAME OF PROXY RESPONDENT] in case it is necessary to clarify any points that come up.**   [If no or asks to be called back later, proceed to end of the survey] [If consent is provided and case/control is 7-18 years old, proceed to 3]

- Participant provided consent on their own behalf (1)
- Parent provided consent for child	(2)
- Participant provided consent for proxy respondent to answer on their own behalf (3)
- No consent was provided (0)

[Only read: if participant is a child younger than 14, and adult is answering on their behalf]

2a. [INSERT CHILD’S NAME] is welcome to stand by or join the call to help answer questions.   [If child joins the call]   2b. Hi [INSERT CHILD’s NAME]. My name is {interviewer name} and I work with the California Department of Public Health. I’m going to ask you some questions about your activities in the past couple of weeks.                          [Proceed to section 3]

**SECTION 3: LAST COVID TEST  (~3 min)** **1.     Great, so to start, I want to ask whether you know your COVID-19 test result from** {test date}**?** *Record whether they know or don’t know their test result*

[If yes, proceed to **section 4**]

[If no, and they are negative, proceed to 2]

[If no, and they are positive, proceed to 3]

*Note that if you are talking to someone who was recently diagnosed with COVID, they might be feeling scared or anxious. It is ok to interject something that sounds human after hearing this, like "Oh I'm so sorry to hear that. I really hope you feel better".*

- Subject knows test result and is positive (1)
- Subject knows test result and is negative (2)
- Subject does NOT know test result and is positive (3)
- Subject does NOT know test result and is negative (4)

Display This Question:

If test_result = 4

**2. Your COVID-19 test result from** {test date} **has come back negative.**   [Proceed to **3**]

**3. Have you ever received a positive COVID-19 test result or been told by a health care provider that you are positive for COVID-19?**

- Yes (1)
- No (2)
- Don't know (3)
- Refuse (4)

Skip To: End of Block If ever_pos = 1

Display This Question:

If test_result = 3

**4.     Your COVID-19 test result from** {test date} **has come back positive. This means you do have coronavirus disease or COVID-19.  In my role with CDPH, I cannot provide you with medical advice. If you need any medical information, please call your healthcare provider. One thing I want to be sure of today is that we have a plan for you to follow up with your healthcare provider, so that they can check on any symptoms you may have and assess your risks. Even if you feel okay now, it is important to have someone you can call if you start feeling sick.** **If you do not have a healthcare provider, you can go to an urgent care facility or the emergency room if you are not getting better or you feel like you are getting worse.**   [Proceed to **section 4**]

[If the person brings up clinical questions or concerns about their positive test] **Thank you for sharing that concern. In my role with CDPH, I am not able to give you medical advice. I do want to be sure that you get the help you need. If you believe you are having a medical emergency, you should call 911. Some warning signs that you should go to the emergency room for are: trouble breathing, bluish lips or face, pain or pressure in the chest that does not go away, new confusion or trouble waking or staying awake, but there are other symptoms too. Otherwise, you should call your healthcare provider.**

**SECTION 4: REASONS FOR TESTING  (~3 min)
1.**     **Next, I am going to ask you some questions about your COVID-19 test. Can you describe to me why you got tested on** {test date} **?** *Interviewers will select check boxes from the respondent based off their response, without prompting them from the following list, and will use a write-in option for any additional reasons for seeking testing. After choosing the best answer from the list, confirm your choice the case/control (ex. "So you got tested for pre or post-travel screening?")*

- I had contact with someone who tested positive (1)
- I had contact with someone who had symptoms, but I do not know if they were confirmed to be positive (2)
- I was told by a public health worker to get tested because I was exposed to a confirmed case (3)
- Someone in my household had contact with someone who tested positive (10)
- A person in my household had contact with someone who had symptoms or suspected they had COVID, but we do not know if they are confirmed to be positive (11)
- I was concerned about symptoms I was experiencing (4)
- Routine screening for my job or school (5)
- Pre or post-travel screening (6)
- Test required for a medical procedure (7)
- I just wanted to see if I was infected (8)
- Testing was required to attend a public event/gathering (15)
- Don't know (12)
- Refuse (13)
- Other: (9) _____________________

**2.     At the time you were tested on** {test date} **were you experiencing any COVID-19 symptoms?**

 [If yes, ask question 3]

[If no, proceed to question 5]

- Yes (1)
- No (0)
- Not Sure (2)
- Refuse (3)

Display This Question:

If any_symptoms = 1

**3.     Can you please list the symptoms you were experiencing on or 14 days prior to your test on** {test date} ? *Interviewers will select the symptoms the respondent indicates they were experiencing. When the respondent is done listing symptoms, the interviewer may prompt, “Are you sure those were all the symptoms you experienced?” and proceed to confirm absence of the 6 most common symptoms (as applicable), in a conversational manner: “No fever, no chills, no muscle pain, no loss of appetite, no shortness of breath, no cough?”*

- Blocked nose (13)
- Chills (5)
- Cough (9)
- Chest pain (23)
- Diarrhea (27)
- Muscle pain (6)
- Fever (4)
- Fatigue (31)
- Headache (12)
- Hoarseness (17)
- Loss of appetite (7)
- Loss of taste (10)
- Loss of smell (11)
- Myalgia (muscle pain) (22)
- Nausea (28)
- Runny nose (14)
- Shortness of breath (8)
- Sneezing (15)
- Sore throat (21)
- Stomach pain (29)
- Sinus pain (24)
- Sweating (19)
- Swollen glands (25)
- Tickle in the throat (20)
- Trouble thinking (30)
- Watery eyes (16)
- Wheezing (26)
- Don't know (35)
- Refuse (36)
- Other (18) __________

**4a.   I am now going to read a list of places you may have sought treatment or advice about a potential COVID-19 infection prior to your test on** {test date}**. After I read the following options, please answer “Yes” or “No”. Ok?**

| **Did you seek care at an in-person appointment with your usual physician or healthcare provider?** | ▼ Yes (1) ... Refuse (4) |
| --- | --- |
| **Did you seek care at a telehealth visit or phone appointment with your usual physician or healthcare provider?** | ▼ Yes (1) ... Refuse (4) |
| **Did you seek care at an in-person visit to an urgent care clinic?** | ▼ Yes (1) ... Refuse (4) |
| **Did you seek care at an in-person visit to a healthcare provider at a retail pharmacy?** | ▼ Yes (1) ... Refuse (4) |
| **Did you visit the emergency room?** | ▼ Yes (1) ... Refuse (4) |
| **Were you admitted to a hospital?** | ▼ Yes (1) ... Refuse (4) |

**4b. And just to follow-up, were there any other forms of health care from which you sought treatment advice at the time you had your test on {test date}?**
 *If so, please specify. If not, leave blank.* 

________________________________________________________________

**5.     In the 14 days prior to your test (between** {14 days prior to test date} **to** {test date}), are you aware of **whether you had known or suspected contact with one or more people who may have tested positive for COVID-19?**

- Yes- contact with one person who was confirmed positive (1)
- Yes- contact with more than one person confirmed positive (2)
- Yes- contact with one person who I suspected was positive (3)
- Yes- contact with more than one person who I suspected was positive (4)
- No known or suspected contact with a positive case (5)
- Not sure (6)
- Refuse (7)

**SECTION 5: CONTACT WITH KNOWN OR SUSPECTED CASE (~8 min)**   [If case indicated they had KNOWN or SUSPECTED Contact , proceed to A] [If case indicated they did NOT have known or suspected contact, proceed to **section 6**]

Display This Question:

If known_exposure = 1

Or known_exposure = 2

Or known_exposure = 3

Or known_exposure = 4

**A.    I’m going to now ask you some questions about the type of contact you had with the person (people) who may have had COVID-19. We are trying to understand sources of exposure and are hopeful that you are willing to answer the questions honestly, knowing that we aren’t looking or expecting any sort of answer.**

Display This Question:

If known_exposure = 1

Or known_exposure = 3

Or known_exposure = 2

Or known_exposure = 4

**1A.     Was the known/ suspected contact someone who lives in your household?**
 *if plural* 
 **Were any of the known/ suspected contact people who lives in your household?**

- Yes (1)
- No (0)
- Don't know (2)
- Refuse (3)

Display This Question:

If known_exposure = 1

Or known_exposure = 2

Or known_exposure = 3

Or known_exposure = 4

2A.     Did the known/ suspected contact occur indoors, outdoors, or both indoors and outdoors?
 *if plural*
 Did the known/ suspected contacts occur indoors, outdoors, or both indoors and outdoors?

- Indoors (1)
- Outdoors (2)
- Both indoors and outdoors (3)
- Unknown (4)
- Refuse (5)

Display This Question:

If known_exposure = 1

Or known_exposure = 2

Or known_exposure = 3

Or known_exposure = 4

**3A.     In the 14 days prior to your test (between** {14 days prior to test date} **to** {test date}**), what are the locations where you may have had contact with this person(s)?**
 *if plural*
 **In the 14 days prior to your test (between** {14 days prior to test date} **to** {test date}**), what are the locations where you may have had contact with these people?**

________________________________________________________________

Display This Question:

If known_exposure = 1

Or known_exposure = 2

Or known_exposure = 3

Or known_exposure = 4

**4A.     I am now going to ask you about different precautions you may or may not have been able to take when you came into contact with the known or suspected positive case(s). Please answer “Yes, No or Not Sure” after each question:**

| **Did you come within 6 feet of this person, indoors?** *if plural:*  **Did you come within 6 feet of any of these people, indoors?** (contact_indoor) | ▼ Yes (1) ... Refuse (4) |
| --- | --- |
| **Did you come within 6 feet of this person, outdoors?** *if plural:*  **Did you come within 6 feet of any of these people, outdoors?** (contact_outdoors) | ▼ Yes (1) ... Refuse (4) |
| **Did you have physical contact with this person, (ie. handshake, hug)?** *if plural:*  **Did you have physical contact with any of these people, (ie. handshake, hug)?** (contact_physical) | ▼ Yes (1) ... Refuse (4) |

Display This Question:

If known_exposure = 1

Or known_exposure = 2

Or known_exposure = 3

Or known_exposure = 4

**5A.     Did you wear a mask the entire time, most of the time, some of the time, or none of the time that you interacted with this person?**
 *if plural:* 
  **Did you wear a mask the entire time, most of the time, some of the time, or none of the time that you interacted with these people?**

- I wore a mask the entire time I interacted with this (these) person(s) (1)
- I wore a mask most of the time I interacted with this (these) person(s) (2)
- I wore a mask some of the time I interacted with this (these) person(s) (3)
- I did not wear a mask during this (these) interaction(s) (4)
- Not sure (5)
- Refuse (6)

Display This Question:

If known_exposure = 1

Or known_exposure = 2

Or known_exposure = 3

Or known_exposure = 4

**6A.     Did the person you had known or suspected contact with wear a mask the all, most, some, or none of the time during your interactions with them?** 
 *if plural:*   
 **Did the people you had known or suspected contact with wear a mask the entire time, most of the time, some of the time, or none of the time when you interacted with them?**

- They wore a mask the entire time we interacted (1)
- They wore a mask most of the time we interacted (2)
- They wore a mask some of the time we interacted (3)
- They did not wear a mask during this interaction (4)
- Not sure (5)
- Refuse (6)

Display This Question:

If known_exposure = 1

Or known_exposure = 2

Or known_exposure = 3

Or known_exposure = 4

**7A. Did you spend more than three consecutive hours with this person in the** 14 days prior to your test (between {14 days prior to test date}  to {test date})
 *if plural:*    
 **Did you spend more than three consecutive hours with these people in the** 14 days prior to your test (between {14 days prior to test date}  to {test date})

- Yes (1)
- No (0)
- Don't know (2)
- Refuse (3)

**SECTION 6: EXPOSURE WITH CONTACT KNOWN OR SUSPECTED CASE- updated (~10 min)**

Next, I want to learn about potential sources of exposure to COVID-19 in the 14 days before your last test: from {14 days prior to test date}  to {test date}. It may help you to pull up a calendar to remember what you were up to over the last two weeks.  **This section can get a bit repetitive, so thanks in advance for your patience with my questions.**    *Only read the following if they did not have known or suspected contact:* We are trying to understand sources of exposure and are hopeful that you are willing to answer the questions honestly, knowing that we aren’t looking or expecting any sort of answer.
 *Read the next statement for all participants:*  I am now going to ask you about a series of locations which you may have visited. After I announce each location, please tell me “Yes, No, or Not sure” to indicate whether you visited that location between {14 days prior to test date}  to {test date}.

**1. First, did you attend a health appointment or health facility (other than where you got tested for COVID-19)?**

- Yes (1)
- No (0)
- Not Sure (2)
- Refuse (3)

Display This Question:

If healthAppt = 1

**1a. How many times did you attend a health appointment or health facility (other than where you got tested for COVID)** between {14 days prior to test date}  to {test date}?

▼ 1 (1) ... Refuse (22)

2. Did you go grocery shopping?

- Yes (1)
- No (0)
- Not Sure (2)
- Refuse (3)

Display This Question:

If groccery = 1

**2a. How many times did you go grocery shopping** between {14 days prior to test date}  to {test date}?

▼ 1 (1) ... Refuse (22)

3. Now I am going to ask you about the times you went to restaurants. Did you go to any restaurants either to pick up take-out or to eat at the restaurant?

- Dine-in (eat at restaurant) only (1)
- Take-out only (2)
- Both dining-in and take-out (5)
- Neither dine-in or take-out (6)
- Not Sure (3)
- Refuse (4)

Display This Question:

If restaurant = 2

Or restaurant = 5

**3a. How many times did you get take-out?**

▼ 0 (1) ... Refuse (23)

Display This Question:

If restaurant = 2

Or restaurant = 5

**3b. Did you ever have to go inside the restaurant to either place or pick up your take-out order?** 
*Interviewer: curb-side pick-up would be considered NOT going inside the restaurant.*

- Yes, I went inside the restaurant either to place or pick-up my order (1)
- No, I did not go inside the restaurant either to place or pick-up my order (2)
- No, I did not go inside the restaurant either to place or pick-up my order, but someone who I went to the restaurant with had to go inside either to place or pick-up the order (5)
- Not Sure (3)
- Refuse (4)

Display This Question:

If restaurant = 1

Or restaurant = 5

**3c. How many times did you eat at an indoor restaurant** between {14 days prior to test date}  to {test date}?
*To clarify, eating at an indoor restaurant means the person spent the majority of their time at the restaurant indoors.*

▼ 0 (23) ... Refuse (22)

Display This Question:

If restaurant = 1

Or restaurant = 5

**3d. How many times did you eat at an outdoor restaurant** between {14 days prior to test date}  to {test date}?
To clarify, eating at an outdoor restaurant means the person spent the majority of their time at the restaurant outdoors.

▼ 0 (23) ... Refuse (22)

[Skip this question if the respondent is under 21- select "underage"]
 4. Did you attend any bars, breweries, or wine bars?

- Yes (1)
- No (0)
- Not Sure (2)
- Refuse (3)
- Underage (4)

Display This Question:

If any_bars = 1

**4a. So did you go to a bar, brewery, or winery?**
 *Select all that apply*

- Brewery (1)
- Bar (2)
- Winery (3)
- Refuse (4)

Display This Question:

If bar_type = 1

**4b. How many times did you** go to a brewery between {14 days prior to test date}  to {test date}?

▼ 1 (1) ... Refuse (22)

Display This Question:

If bar_type = 1

**4c. When you went to a (those) brewery(ies), did you spend most of your time indoors, outdoors, or both indoors and outdoors?**

- Indoors (1)
- Outdoors (2)
- Both indoors and outdoors (3)
- Don't know (4)
- Refuse (5)

Display This Question:

If bar_type = 2

**4d. How many times did go to a bar** between {14 days prior to test date}  to {test date}?

▼ 1 (1) ... Refuse (22)

Display This Question:

If bar_type = 2

**4e. When you went to a (those) bar(s), did you spend most of your time indoors, outdoors, or both indoors and outdoors?**

- Indoors (1)
- Outdoors (2)
- Both indoors and outdoors (3)
- Don't know (4)
- Refuse (5)

Display This Question:

If bar_type = 3

**4f. How many times did you** attend a winery between {14 days prior to test date}  to {test date}?

▼ 1 (1) ... Refuse (22)

Display This Question:

If bar_type = 3

4g. When you went to a (those) winery(ies), did you spend most of your time indoors, outdoors, or both indoors and outdoors?

- Indoors (1)
- Outdoors (2)
- Both indoors and outdoors (3)
- Don't know (4)
- Refuse (5)

**5. Did you ever visit a coffee or tea shop?**

- Yes (1)
- No (0)
- Don't know (2)
- Refuse (3)

Display This Question:

If coffee = 1

**5a. How many times did you visit a coffee or tea shop between {14 days prior to test date}  to {test date}?**

▼ 0 (1) ... Refuse (23)

Display This Question:

If coffee = 1

**5b. When you (typically) visited the coffee or tea shop(s), did you have to go inside to place your order?**

- I went inside to place the order (1)
- I typically placed the order outside, or I typically placed the order remotely (ex. via app, web portal, phone order) (2)
- Don't know (3)
- Refuse (4)

Display This Question:

If coffee = 1

**5c. When you visited the coffee or tea shop(s), did you (typically) consume your beverage inside the shop, outside the shop, or did you just pick-up the beverage for take-away?**

- Consumed inside the shop (1)
- Consumed outside the shop (ex. restaurant set up outdoor tables/ chairs and I drank/ ate at those tables) (2)
- Got beverage for take-away (3)
- Don't know (4)
- Refuse (5)

6. Did you go retail shopping?

- Yes (1)
- No (0)
- Not Sure (2)
- Refuse (3)

Display This Question:

If retail_shop = 1

**6a. And did you go indoor or outdoor retail shopping?**  *Indoor retail shopping = spend the majority of your time inside (ex. Target, Walmart)* *Outdoor retail shopping = spend the majority of your time outside (ex. flea market, street festival, farmers market)* *Indoor and outdoor retail shopping = Shopping at a retail site that has both an outdoor and indoor venue (ex. Home Depot, or a boutique that has set up clothing outside, but also has items inside to browse)*

- Indoor (1)
- Outdoor (2)
- Both indoor and outdoor (3)
- Don't know (5)
- Refuse (4)

Display This Question:

If retail_inside = 1

**6b. How many times did you** go indoor retail shopping between {14 days prior to test date}  to {test date}?

▼ 1 (1) ... Refuse (22)

Display This Question:

If retail_inside = 2

**6c. How many times did you** go outdoor retail shopping between {14 days prior to test date}  to {test date}?

▼ 1 (1) ... Refuse (22)

Display This Question:

If retail_inside = 3

**6d. How many times did you** go retail shopping that was both indoors and outdoors between {14 days prior to test date}  to {test date}?
Ex. *Shopping at a retail site that has both an outdoor and indoor venue (ex. Home Depot, a boutique that has set up clothing outside, but also has items inside to browse)*

▼ 1 (1) ... Refuse (22)

gym 7. Did you exercise at a gym?

- Yes (1)
- No (0)
- Don't know (2)
- Refuse (3)

Display This Question:

If gym = 1

**7a. And was this an indoor or an outdoor gym?**

- Indoor (1)
- Outdoor (2)
- Both indoor and outdoor (3)
- Don't know (5)
- Refuse (4)

Display This Question:

If gym_inside = 1

**7b. How many times did you** exercise at an indoor gym between {14 days prior to test date}  to {test date})?

▼ 1 (1) ... Refuse (22)

Display This Question:

If gym_inside = 2

**7c. How many times did you** exercise at an outdoor gym between {14 days prior to test date}  to {test date})?

▼ 1 (1) ... Refuse (22)

Display This Question:

If gym_inside = 3

**7d. How many times did you** exercise at a gym that was both indoors and outdoors between {14 days prior to test date}  to {test date})?

▼ 1 (1) ... Refuse (22)

8. Did you participate in a group recreational sport like tennis, soccer, basketball, or swimming?

- Yes (1)
- No (0)
- Don't know (2)
- Refuse (3)

Display This Question:

If sport = 1

**8a. How many times did you do a group recreational sport?**

▼ 1 (1) ... Refuse (22)

9. Did you ever leave your house to go for a walk, run, hike, or bike ride outside?

- Yes (1)
- No (0)
- Not Sure (2)
- Refuse (3)

Display This Question:

If walk = 1

**9a. How many times did you walk, run, hike, or ride a bike outdoors between {14 days prior to test date}  to {test date}?**

▼ 1 (1) ... Refuse (22)

Display This Question:

If walk = 1

**9b. Did you ever hike, run, walk, or bike with anyone outside your household?**

- No, I always hiked, ran, walked, or biked by myself (1)
- No, but I sometimes/ always hiked, ran, walked, or biked with other people who live in my household (5)
- Yes, I hiked, ran, walked, or biked with someone who doesn't live in my household (2)
- Don't know (3)
- Refuse (4)

10. Did you ride public transit?

- Yes (1)
- No (0)
- Don't know (2)
- Refuse (3)

Display This Question:

If public_transit = 1

**10a. How many times did you ride public transit** between {14 days prior to test date}  to {test date}?

▼ 1 (1) ... Refuse (22)

Display This Question:

If public_transit = 1

**10b. What type of public transit did you ride between** {14 days prior to test date}  to {test date}?
*Interviewers- select all that apply*

- Public Bus (1)
- Subway (2)
- Commuter train (light rail) (25)
- Ferries or water taxis (23)
- Trolley (24)
- Other (26) ______________
- Refuse (27)
- Don't know (28)

11. Did you use a ride share service (like Taxi, Uber, Lyft, or carpool with individuals who are not members of your household)?

- Yes (1)
- No (0)
- Not Sure (2)
- Refuse (3)

Display This Question:

If ride_share = 1

**11a. How many times did you use a ride-share service** between {14 days prior to test date}  to {test date}?

▼ 1 (1) ... Refuse (22)

12. Did you fly on a plane?

- Yes (1)
- No (0)
- Don't know (2)
- Refuse (3)

Display This Question:

If plane = 1

**12a. How many times did you fly on a plane** between {14 days prior to test date}  to {test date}?

▼ 1 (1) ... Refuse (22)

Display This Question:

If plane = 1

**12b. Did you fly domestically or internationally?**

- Domestic (1)
- International (2)
- Both (23)
- Don't know (3)
- Refuse (4)

Display This Question:

If plane = 1

**12c. If you had to estimate, what was your total travel time between arrival at your departure airport and exit at your arrival airport?**
*Interviewers: prompt to make sure they understand you are not just asking about travel time on the plane, but the entire travel time in the airport*

- under 3 hours (1)
- 4-5 hours (2)
- 6-7 hours (3)
- 8-10 hours (4)
- 10-12 hours (23)
- 12-15 hours (24)
- 15-20 hours (26)
- 20-24 hours (27)
- over 24 hours (28)
- Don't know (29)
- Refuse (30)

13. Did you attend a parade, rally, march, or protest?

- Yes (1)
- No (0)
- Don't know (2)
- Refuse (3)

Display This Question:

If march = 1

**13a. How many times did you** attend a parade, rally, march, or protest between {14 days prior to test date}  to {test date}?

▼ 1 (1) ... Refuse (22)

14. Did you receive services at a salon or barber shop?

- Yes (1)
- No (0)
- Don't know (2)
- Refuse (3)

Display This Question:

If salon = 1

**14a. How many times did you receive services at a salon or barber** between {14 days prior to test date}  to {test date}?

▼ 1 (1) ... Refuse (22)

Display This Question:

If salon = 1

**14b. Was the salon or barbershop that you received services at indoors, outdoors, or both indoors and outdoors?**

- Indoors (1)
- Outdoors (2)
- Both indoors and outdoors (3)
- Don't know (4)
- Refuse (5)

15. Did you attend an indoor movie theater?

- Yes (1)
- No (0)
- Don't know (2)
- Refuse (3)

Display This Question:

If movie = 1

**15a. How many times did you** attend an indoor movie theater between {14 days prior to test date}  to {test date}?  

▼ 1 (1) ... Refuse (22)

16. Did you attend a worship service?

- Yes (1)
- No (0)
- Don't know (2)
- Refuse (3)

Display This Question:

If worship = 1

**16a. Was the worship service indoor or outdoors?**

- Indoors (1)
- Outdoors (2)
- Both Indoors and outdoors (3)
- Don't know (5)
- Refuse (4)

Display This Question:

If worship_inside = 1

**16b. How many times did you** attend an indoor worship service between {14 days prior to test date}  to {test date}?

▼ 1 (1) ... Refuse (22)

Display This Question:

If worship_inside = 2

**16c. How many times did you** attend an outdoor worship service between {14 days prior to test date}  to {test date}?

▼ 1 (1) ... Refuse (22)

Display This Question:

If worship_inside = 3

**16d. How many times did you** attend a worship service that was both indoors and outdoors between {14 days prior to test date}  to {test date}?

▼ 1 (1) ... Refuse (22)

17. Did you visit or stay at a school, daycare, or preschool?

- Yes (1)
- No (0)
- Don't know (2)
- Refuse (3)

Display This Question:

If school = 1

**17a. How many times did you** visit or stay at a school, daycare, or preschool between {14 days prior to test date}  to {test date}?

▼ 1 (1) ... Refuse (22)

Display This Question:

If school = 1

17b. Was the school or daycare public or private?

- Public (1)
- Private (2)
- Other (5) ________________________________________________
- Not Sure (3)
- Refuse (4)

17c. Did you visit any theme parks or amusement parks?

- Yes (1)
- No (0)
- Don't know (2)
- Refuse (3)

**I am now going to ask you some questions about face coverings.**

**18.** Between {14 days prior to test date}  to {test date}, at all of the public, indoor places we just discussed, **did you wear a face mask all, most, some, or none of the time?**  *Interviewer: note that we will ask about social gatherings and mask usage separately.*

- I wore a face mask all of the time	(1)
- I wore a face mask most of the time	(2)
- I wore a face mask some of the time (3)
- I never wore a face mask in indoor places (4)
- Refuse (5)
- Don't know (6)
- I did not go inside any indoor places other than my home (7)

**19. B**etween {14 days prior to test date}  to {test date}, a**t all of the public, indoor places we discussed earlier, did people you came within 6 feet of** wear a face mask all, most, some, or none of the time?

- They wore a face mask all of the time	(1)
- They wore a face mask most of the time	(2)
- They wore a face mask some of the time	(3)
- They never wore a face mask in indoor places (4)
- Refuse (5)
- Don't know (6)
- I did not go inside any indoor places other than my home (7)
- I was not in contact with any people outside my household in indoor places (8)

**20.** Between {14 days prior to test date}  to {test date}, at all of the public, outdoor places we discussed earlier, did you wear a face mask all, most, some, or none of the time?

- I wore a face mask all of the time	(1)
- I wore a face mask most of the time	(2)
- I wore a face mask some of the time	(3)
- I never wore a face mask in outdoor places (4)
- Refuse (5)
- Don't know (6)
- I did not go inside any outdoor places other than my home (7)

**21.** Between {14 days prior to test date}  to {test date}, at all of the public, outdoor places we discussed earlier, did people you came within 6 feet of wear a face mask all, most, some, or none of the time?

- They wore a face mask all of the time	(1)
- They wore a face mask most of the time	(2)
- They wore a face mask some of the time	(3)
- They never wore a face mask in outdoor places (4)
- Refuse (5)
- Don't know (6)
- I have not left my home to go outside in the past 14 days (7)
- I was not in contact with any people outside my household in outdoor places (8)

**22. Ok thank you. Now I want to transition into asking you some questions about social gatherings. These would include any informal gatherings with friends or family who are NOT members of your household at locations other than those we already discussed, including your own home. Did you attend any social gatherings** between {14 days prior to test date}  to {test date}? 
 *Interviewer: note that our definition of social gatherings is mixing with people who don't otherwise live in your household. If someone had a longer-term family* together *(ie. traveled to visit relatives, but stayed for multiple days, count this as ONE event). 

NOTE: if they say no, please confirm that they did not have any other gatherings with individuals outside their household in any settings*

- Yes (1)
- No (0)
- Not Sure (2)
- Refuse (3)

Display This Question:

If any_gathering = 1

23a. When you attended social gatherings, were they indoor-only social gatherings, outdoor only social gatherings, or social gatherings that were held both indoors and outdoors?  *Interviewer: select all that apply. 
If its confusing, ask the interviewer "what kind of social gatherings did you attend?" and try to bucket them into events that were exclusively outdoors, exclusively indoors, or events held both inside and outside.*    *An outdoor only gathering means the person spent the majority of their time outside*  *An indoor only gathering means the person spent the majority of their time*  *A gathering that was "both indoors and outdoors" means the participant was both inside and outside during the social gathering (ex. Sandy had some friends over for dinner and they ate outside on the patio, and then watched a movie in their living room together)*

- An indoor only gathering (1)
- An outdoor only gathering (2)
- A gathering that was both indoors and outdoors (5)
- Don't know (3)
- Refuse (4)

Display This Question:

If social_inside = 1

**23b. How many indoor social gatherings did you attend** between {14 days prior to test date}  to {test date}?

▼ 1 (1) ... Refuse (22)

Display This Question:

If social_inside = 1

**23c. On average, about how many total people attended this (these) indoor gathering(s)?**

▼ 1 (1) ... Refuse (22)

Display This Question:

If social_inside = 1

**23d. Did you eat or drink during this (any of these) indoor gathering(s)?**

- Yes (1)
- No (0)
- Don't know (2)
- Refuse (3)

Display This Question:

If social_inside = 1

**23e. When you attended this (these) indoor gathering(s), did you wear a face mask all, most, some or none of the time?**

- I wore a face mask all of the time (1)
- I wore a face mask most of the time (2)
- I wore a face mask some of the time (6)
- I wore a face mask none of the time (5)
- Don't know (3)
- Refuse (4)

Display This Question:

If social_inside = 2

**23f. How many outdoor social gatherings did you attend** between {14 days prior to test date}  to {test date}?

▼ 1 (1) ... Refuse (22)

Display This Question:

If social_inside = 2

**23g. On average, about how many people attended this (these) outdoor gathering(s)?**

▼ 1 (1) ... Refuse (22)

Display This Question:

If social_inside = 2

**23h. Did you eat or drink during this (any of these) outdoor gathering(s)?**

- Yes (1)
- No (0)
- Don't know (2)
- Refuse (3)

Display This Question:

If social_inside = 2

**23i. When you attended this (these) outdoor gathering(s), did you wear a face mask all, most, some or none of the time?**

- I wore a face mask all of the time (1)
- I wore a face mask most of the time (2)
- I wore a face mask some of the time (6)
- I wore a face mask none of the time (5)
- Don't know (3)
- Refuse (4)

Display This Question:

If social_inside = 5

**23j. How many social gatherings did you attend that were both indoor and outdoors** between {14 days prior to test date}  to {test date}?
 *Interviewer: an example of this type of social gathering would me "Sandy had friends over at her patio for dinner, and when it got cold they went inside"*

▼ 1 (1) ... Refuse (22)

Display This Question:

If social_inside = 5

**23k. On average, about how many total people attended this (these) indoor and outdoor gathering(s)?**

▼ 1 (1) ... Refuse (22)

Display This Question:

If social_inside = 5

**23l. Did you eat or drink during this (any of these) indoor and outdoor gathering(s)?**

- Yes (1)
- No (0)
- Don't know (2)
- Refuse (3)

Display This Question:

If social_inside = 5

**23m. When you attended this (these) indoor and outdoor gathering(s), did you wear a face mask all, most, some or none of the time?**

- I wore a face mask all of the time (1)
- I wore a face mask most of the time (2)
- I wore a face mask some of the time (6)
- I wore a face mask none of the time (5)
- Don't know (3)
- Refuse (4)

**24. And last question for this section: did you attend any other kind of event where there were 5 or more people who are not in your household in attendance between** {14 days prior to test date}  to {test date}?
 *If necessary, prompt with options like a sporting event, concert, festival (etc)*

- Yes (1)
- No (0)
- Not Sure (2)
- Refuse (3)

Display This Question:

If other_event = 1

**24a. What was this event?**

________________________________________________________________

**SECTION 7: OCCUPATION (~1 min)**

**1.     I am now going to ask you some questions about your occupation.** Between {14 days prior to test date}  to {test date}, did you **you attend work, school, or volunteering commitments exclusively at home, both at home and "in-person", or exclusively "in-person"?**

- I worked, studied, and/or volunteered exclusively at home (1)
- I attended work, school, and/or volunteered both "in-person" and at home (4)
- I attended work, school, and/or volunteered exclusively “in-person” (2)
- I was not working, in school, or in a volunteer position (3)
- Don't know (5)
- Refuse (6)

Display This Question:

If work_loc = 1

Or work_loc = 4

Or work_loc = 2

[If the respondent is a student, skip question and just record "student"]

**2. Can you tell me what your job is?**
 *Record the free-response of the subject.*
[If they attend work, school, or volunteering commitments in person or both at home & in person, proceed to question 3, otherwise proceed to **Section 8**]

________________________________________________________________

Display This Question:

If work_loc = 2

Or work_loc = 4

*Tailor these next two questions based off of whether the person works, is in school, or volunteers*

**3a. Do you come into close contact (within 6 feet) of more than 10 people per day at work/school/volunteering?**

- Yes (1)
- No (0)
- Don't know (2)
- Refuse (3)

Display This Question:

If work_loc = 2

Or work_loc = 4

**3b. Do you primarily attend work/school/volunteering indoors, outdoor, or both indoors and outdoors?**

- Indoors (1)
- Outdoors (2)
- Both indoors and outdoors (3)
- Don't know (4)
- Refuse (5)

**SECTION 8: VACCINATION (~2 min)**   **I am now going to ask you some questions about the COVID-19 vaccine.**

**1. Do you have any conditions that might place you at higher risk for COVID-19?** *Interviewers may prompt with examples such as diabetes, high blood pressure, obesity, being immunocompromised if requested. Select the conditions from the list below or click "No conditions"*

- No conditions (30)
- Lung conditions: COPD (6)
- Lung conditions: lung cancer (7)
- Lung conditions: Cystic fibrosis (8)
- Lung conditions: moderate to severe asthma (9)
- Lung conditions: Pulmonary fibrosis (10)
- Heart disease (11)
- High blood pressure (23)
- Obesity (12)
- Overweight (13)
- Diabetes (14)
- Weakened immune system: organ transplant (15)
- Weakened immune system: cancer treatment (16)
- Weakened immune system: bone marrow transplant (17))
- Weakened immune system: HIV/AIDS (18)
- Blood disorders: sickle cell anemia (19)
- Blood disorders: thalassemia (20)
- Chronic kidney disease (21)
- Chronic liver disease (22)
- Pregnant: first trimester (27)
- Pregnant: second trimester (28)
- Pregnant: third trimester (29)
- Don't know (25)
- Refuse (26)
- Other (24) _______________

**2.     Have you now or previously had access to a COVID-19 vaccine on the premises of your work or school?**

- Yes (1)
- No I do not currently nor have I ever had access to the vaccine at work/ school (0)
- No I am not currently in work or school (4)
- Don't know (2)
- Refuse (3)

**3.     Have you received any doses of a COVID-19 vaccine?**

- Yes (1)
- No (0)
- Don't know (2)
- Refuse (3)

Display This Question:

If vx = 0

Or vx = 2

3a.     Do you plan to receive the COVID-19 vaccine when it becomes **available** to you?

- Yes (1)
- No (2)
- Don't know (3)
- Refuse (4)

Display This Question:

If vx_hesistancy = 2

Or vx_hesistancy = 3

3b. Can you describe to me why you are not planning to receive the COVID-19 vaccine?
*Interviewers: select all that apply*

- I have a contraindicated medical condition (e.g. severe allergic reaction to a component of the vaccine) (12)
- I have had negative reactions to vaccines in the past (21)
- I do not think the COVID-19 vaccine is safe, but I generally get other vaccines (1)
- I do not think most vaccines, including the COVID-19 vaccines, are safe (11)
- I am concerned about the long-term effects of COVID-19 vaccines (6)
- I am concerned about side effects (2)
- I am pregnant or breastfeeding and do not want to get the vaccine yet (15)
- I do not trust the government (5)
- I do not trust medical professionals or public health agencies (17)
- I already had an infection with COVID-19, so I do not need the vaccine (8)
- I am not at high risk, so I don't think I need the vaccine (10)
- I do not think the COVID-19 vaccine is important to receive (7)
- I am afraid the vaccine will give me COVID-19 (16)
- I am afraid of needles (18)
- I am worried about the cost of the vaccine (19)
- I would only get it if it was required by my job or school (20)
- I am not able to take time off of work to travel to a vaccine site (22)
- The vaccine sites are located too far from my home (23)
- I am under 18/ waiting for trial results for vaccine safety in children (24)
- I object to the vaccine for religious reasons (25)
- I just don't know enough about COVID-19 vaccines (26)
- Depends on what brand of vaccine I am offered (28)
- Waiting to see more research (29)
- It is my personal choice (30)
- Other: (9) __________________
- Don't know (3)
- Refuse (4)

Display This Question:

If vx = 1

**4. How many doses of the COVID-19 vaccine have you received?**

- 1 (1)
- 2 (2)
- 3 (5)
- Not sure (3)
- Refuse (4)

Display This Question:

If vx = 1

**5. Do you have a vaccine card on hand from when you got the COVID-19 vaccine?** *if yes, ask them to get their vaccine card. If no, ask them to do their best remembering and try pulling up a calendar to help them remember.*

- Yes (1)
- No (2)
- No vaccine card, but referenced calendar/ email/ other source (5)
- Don't know (3)
- Refuse (4)

Display This Question:

If vx = 1

**6. What date did you receive your FIRST dose?** *Enter date as MM/DD/YYYY or write "UNKNOWN" or "REFUSED"* 

________________________________________________________________

Display This Question:

If vx = 1

And doses = 2

Or doses = 5

**7. What date did you receive your SECOND dose?** *Enter date as MM/DD/YYYY or write "UNKNOWN" or "REFUSED"
INTERVIEWER: please do a sanity check to make sure the dates they are giving you seem plausible and prompt with something like: "Just to confirm, you received your first dose on [date], and then 10 days later on [date] you received your second dose?”*
- Pfizer second dose should be 21+ days after the first dose
- Moderna second dose should be 28+ days after the first dose
- There should not be a second dose of Johnson & Johnson 
- If someone received two different vaccine products, it is probably reasonable they got their second dose 21+ days after the first 

________________________________________________________________

Display This Question:

If vx = 1

**8. What product did you receive for your FIRST dose?** *Interviewer: if it seems like the person doesn't know what product they received, try to prompt them with the names listed on this page to help them remember*

- Pfizer/BioNTech (1)
- Moderna (2)
- Johnson & Johnson (3)
- Novavax (6)
- Don't know (9)
- Refuse (5)
- Other: (8) ________________________________________________

Display This Question:

If vx = 1

And doses = 2

Or doses = 5

**9. What product did you receive for your SECOND dose?** *Interviewer: if it seems like the person doesn't know what product they received, try to prompt them with the names listed on this page to help them remember*

- Pfizer/BioNTech (1)
- Moderna (2)
- Johnson & Johnson (3)
- Novavax (7)
- Refuse (5)
- Don't know (6)
- Other: (4) ________________________________________________

Display This Question:

If vx = 1

**9a. Have you received any booster doses of a COVID-19 vaccine product?**    *Note to interviewer: booster doses are currently (8/17) recommended for people with immunocompromised conditions and are recommended four weeks after completion of the second dose in a mRNA vaccine series. People who received J&J are not yet recommended to get a booster dose. https://www.cdc.gov/coronavirus/2019-ncov/vaccines/recommendations/immuno.html*

- Yes (5)
- No (6)
- Not sure (7)
- Refuse (8)

Display This Question:

If vx = 1

And any_booster = 5

**9b. What date did you receive your BOOSTER dose?**  *Enter date as MM/DD/YYYY or write "UNKNOWN" or "REFUSED"*

________________________________________________________________

Display This Question:

If vx = 1

And any_booster = 5

**9c. What product did you receive for your BOOSTER dose?**  *Interviewer: if it seems like the person doesn't know what product they received, try to prompt them with the names listed on this page to help them remember*

- Pfizer/BioNTech (1)
- Moderna (2)
- Johnson & Johnson (3)
- Novavax (7)
- Refuse (5)
- Don't know (6)
- Other: (4) ________________________________________________

Display This Question:

If vx = 1

**10. Where did you get your COVID-19 vaccine?** 
*Note to interviewer: if they received a primary and booster series, select the location(s) of both doses if it varies*

- Mass vaccination site (1)
- Hospital (0)
- Nursing home or assisted living facility (4)
- Private physician office (10)
- At a retail pharmacy (eg. Walgreens) (8)
- Mobile vaccination site/ community outreach vaccination program (11)
- At my work (select Hospital/ Nursing home if they work at the hospital/ nursing home and got the vaccine there) (6)
- At a school (7)
- At a retail shop (eg. Walmart) (9)
- Correctional facility (5)
- Urgent care or community clinic (13)
- County or state health department (14)
- Refuse (3)
- Don't know (2)
- Other (12) ________________________________________________

Display This Question:

If vx = 1

**11. At the time you received the vaccine were you required to receive it to attend work or school?**

- Yes (1)
- No (0)
- Not sure (4)
- Refuse (5)

**SECTION 9: DEMOGRAPHICS  (~5 min)**   **I just have a few more questions. Again, anything you share with me is confidential and protected by California’s strict privacy laws. The information we collect about you will assist the health department in their COVID-19 response.**

**1.    First, I’m going to ask you some general questions about COVID-19. In the two weeks prior to getting your COVID-19 test, how worried did you feel about getting COVID-19? Would you say you felt:**

- **Very worried** (1)
- **Somewhat worried** (2)
- **Neutral** (3)
- **Not worried at all** (4)
- Don't know (5)
- Refuse (6)

**2.    Since the beginning of the pandemic, there have been a lot of recommendations on behaviors that can reduce the risk of COVID-19 including avoiding large crowds, travel, and maintaining 6 feet of distance in public places. Would you say that you strongly agree, agree, are neutral, disagree, or strongly disagree that these measures reduce the risk of COVID-19?**

- Strongly agree (1)
- Agree (2)
- Neutral (3)
- Disagree (4)
- Strongly Disagree (7)
- Don't know (5)
- Refuse (6)

**3.    Another recommendation to reduce the spread of COVID-19 is wearing face masks. Would you say that you strongly agree, agree, are neutral, disagree, or strongly disagree that face masks reduce the risk of COVID-19?**

- Strongly agree (1)
- Agree (2)
- Neutral (3)
- Disagree (4)
- Strongly Disagree (7)
- Don't know (5)
- Refuse (6)

**Last, there's just a couple questions about demographics and a few about housing left.**

**4.   So do you mind sharing how old you are?**
 *Please select "REFUSE" if they decline to answer*

▼ under 12 mo (4) ... Refuse (105)

**Next, can you describe your ethnic background?**
*Leave the question blank if they refuse to answer* *Interviewer: use the box to select and write-in the ethnic background the participant identifies as if they provide you more information. You may select all that apply. 
Examples of write-in text:*  Black: African-American, Caribbean (Jamaican, Haitian), African (Nigerian, Ethiopian, Somali, Ghanian, South African, Kenyan) 
Asian: Chinese, Filipino, Vietnamese, Korean, Japanese, Pakistani, Cambodian, Asian Indian 
Hispanic, Latino, or Spanish: Mexican, Puerto Rican, Cuban, Guatemalan, Ecuadorian, Venezuelan, Peruvian, Dominican 
American Indian or Alaska Native: Navajo nation, Blackfeet tribe
Native Hawaiian or Pacific Islander**:** Native Hawaiian, Somoan, Tongan, Fijian 
Middle Eastern: Persian, Arab, Egyptian, Syrian, Israeli, Middle Eastern, Iranian, Moroccan, Palestinian
White: English, Irish, Italian, Eastern European

- White (1) _____________
- Black (2) ______________
- Hispanic, Latino, or Spanish (3) ____________
- Asian (4) ____________
- Native Hawaiian or Pacific Islander (5) ____________
- American Indian or Alaska Native (6) _____________
- Middle Eastern (7) ___________

**5. What is your sex?**
 *Please select "REFUSE" if they decline to answer.*

- Woman (5)
- Man (6)
- Non-binary (7)
- Prefer to self-describe, below: (8) ___________________
- Refuse (9)
- Don't know (10)

**6a. What is the zip-code of your home address?**
*If they are unwilling or don't know, please write "REFUSE". Encrypt the zip-code following SOP instructions, and paste the encrypted zip code below:*
 
***NOTE: If RSA website is down, follow temporary instructions on*** [SOP](https://docs.google.com/document/d/1Hd3hs2gbQ28q90r6jGUzzS7o45MHfbvIqwZx5TIzobI/edit?ts=602b7883) ***to record address and zip code info***

**6b. What is your home address?**
 If they are unwilling or don't know, please write "REFUSE". You can preface with: "**We use the information only for geospatial analysis and will not send anything to your address. We encrypt the information so that it is secure**." Encrypt the home address following SOP instructions, and paste the encrypted zip code below:
 
***NOTE: If RSA website is down, follow temporary instructions on*** [SOP](https://docs.google.com/document/d/1Hd3hs2gbQ28q90r6jGUzzS7o45MHfbvIqwZx5TIzobI/edit?ts=602b7883) ***to record address and zip code info***

**7. Which of the following best describes your living arrangement:**

- **Private home (1)**
- **Apartment or condominium (2)**
- **Skilled nursing facility (3)**
- **College or university student housing (4)**
- **Military quarters (5)**
- **Emergency or transitional shelter (6)**
- **Other (or indicate here if unhoused): (7) ________________**
- Don't know (8)
- Refuse (9)

**8.     How many people (other than yourself) live in your household?**

▼ 0 (12) ... REFUSE (11)

**9.     How many bedrooms do you have in your household?**

▼ 0 (studio) (12) ... REFUSE (11)

**10. Are there any children under 18 living in your household?**

- Yes (1)
- No (0)
- Refuse (3)
- Don't know (2)

Display This Question:

If children = 1

**11. Are any of your children under 18 attending in-person instruction, school, or daycare?**

- Yes (1)
- No (0)
- Refuse (3)
- Don't know (2)

**12. Does anyone visit your home on a regular basis like a cleaning service or babysitter?**

- Yes (please write-in the "service" who visits the home below) (1) ________________
- No (0)
- Don't know (2)
- Refuse (3)

If you are talking to a child aged 14-17, at this point you can end the interview with the child and ask to speak with their parent/guardian. When you get back on the phone with the parent or guardian, you can say something like **["Hi again, thank you so much for letting me speak with your child, it was extremely helpful. We are wrapping up the survey with some demographic questions and my last question that I didn't want your child to have to answer was whether you are willing to share your total household income?"]**

**13. And lastly, information about your total household income is extremely helpful for us. If you don't mind sharing, is your total household income...**
If they ask, tell them to answer on behalf of everyone you share finances with. 

- **Under $50,000** (4)
- **$50,000 to $100,000** (5)
- **$100,000 to $150,000** (6)
- **Over $150,000** (7)
- Not sure (8)
- Refuse (9)

Display This Question:

If test_result = 1

Or test_result = 3

**Thank you for participating in this survey. You may be contacted by another staff member at the health department to check in on you. They will ask you questions about your health and well-being to make sure you’re ok. Before I end the call, do you have any questions for me about the survey?**

Display This Question:

If test_result = 2

Or test_result = 4

**Thank you for participating in our survey. We appreciate your time. Before I end the call, do you have any questions for me about the survey?**

End of Block: Section 1: Introduction

Start of Block: Final block

Display This Question:

If consent = 0

**2. No problem. Before you hang up, do you mind quickly sharing why you are unable or unwilling to complete this call today?**

- Not a good time/ I don't have time right now (1)
- Not interested (2)
- Refuse to answer (4)
- Hung up before you could ask this question (5)
- Other: (3) ________________________________________________

Display This Question:

If ever_pos = 1

[Read if the person is has previously tested COVID positive] 
 Thanks for letting me know. Unfortunately, **because** of that previous positive, I am unable to continue with the interview. But I really **appreciate** your willingness to participate in our survey. Thank you so much for your time! 
[Make sure you click that the survey was terminated early because the case/control had previously tested positive]

Display This Question:

If understand = 4

No worries. Thank you for your time and I hope you have a nice day.  

**If the survey was completed, did you remember to encrypt both the zip code and the address? Remember to follow instructions outlined in the** [SOP](https://docs.google.com/document/d/1Hd3hs2gbQ28q90r6jGUzzS7o45MHfbvIqwZx5TIzobI/edit?ts=602b7883) **to 1) copy and paste a new public key and 2) select (** 'RSA/NONE/OAEPWithSHA1AndMGF1Padding.' ) under RSA ciphers. 
  Interviewer: Please select the call outcome

- Survey respondent consented and completed the entire survey (8)
- Survey respondent consented and completed a partial survey (choose this for every call that was terminated after obtaining consent, including cases/controls who previously tested positive) (1)
- Survey respondent did NOT consent (choose this if they explicitly did not consent, hung up before you could ask for consent, or did not understand the purpose of the survey) (13)

Display This Question:

If call_outcome = 1

Interviewer: why was the survey terminated AFTER consent was already provided?

- Subject was a case/control who had previously tested positive (5)
- Subject just terminated the call after consent was provided or call dropped (4)
- Subject terminated the call early for a specific reason (write-in the reason) (6) ________________________________________________

Display This Question:

If call_outcome = 13

Interviewer: why was the survey terminated early?

- Subject did not understand information due to language barrier (1)
- Subject was not mentally-fit to respond to survey questions (2)
- Subject did not provide consent (3)
- Call just dropped (5)
- Other reason: (4) ________________________________________________

OPTIONAL: Interviewers may add any interview notes that you took during the interview to this free text box before submitting the survey.

________________________________________________________________

Interviewer: select this box if the survey was completed in spanish

- I conducted the survey in spanish (1)

Did you encrypt both the zip code and the address of the participant if they provided it?

- Yes- I have verified that both the zip code and address are appropriately encrypted (1)

End of Block: Final block

**Item S2.** Interviewer experience survey.

C4 study reflection survey

Survey Flow

Section 1: Introduction (1 Question)

Section 2: Impact and/or mental health (8 Questions)

Section 3: Weekly meetings (2 Questions)

Section 4: Messaging platform (2 Questions)

Section 5: Overall feedback (5 Questions)

All questions are optional to complete the survey. Survey is anonymous by default, unless participants explicitly choose to provide a name.

Section 1: Introduction

**SECTION 1**

Hello! As the study comes to a close, we are looking to learn from the experience of being a part of this study. Results are anonymous unless you choose to include your name below.

As interviewers, you have tremendous insight about the implementation of this study, so if you have the time to give detailed or thoughtful responses, it would be extremely helpful.

A response by Monday, July 11 would be appreciated. Thank you!!

We will be including a summary of the key insights you share in the upcoming implementation paper to give a qualitative perspective to what implementing this study was like for interviewers. The hope is that we can share with future researchers of similar studies what to anticipate, what additional preparations might be improved upon to support interviewers better or make the study more effective etc.

You can choose to speak towards any of the themes (you can skip around sections), but are not required to answer all questions (although that would be appreciated!) . For each theme you choose to respond to, please select whether or not it would be alright to include any quotes of what you wrote in the paper. Quotes will be anonymous.

1. **OPTIONAL: If you would like to be non-anonymous for any reason, you can provide a name here. Also, if you would like for us to follow up with you about any concerns, please indicate that as well.** (Long answer text)

Section 2: Impact and/or mental health

**SECTION 2**
This section asks questions that may or may not bring up uncomfortable memories. If you feel that it may be too difficult to discuss the topics mentioned, please skip ahead to the next section.

1. **Are there any interviewing experiences that impacted you strongly? Please describe them and their impact on you.** (Long answer text)
2. **How did the mental health of the people you interviewed impact you during calls?** (Long answer text)
3. **How often did the people you interviewed ask you for resources or confide something concerning that compelled you to look for /share resources?** (Multiple choice)
   - Almost never
   - Occasionally
   - Often
   - Almost all the time
4. **What resources did they ask for or ? Select all that apply** (Check boxes)
   - Food security related
   - Housing security related
   - Healthcare access related
   - COVID-19 vaccine or treatment related
   - Safety related
   - Mental health related
   - Financial related (eg. asking for money)
   - Other
5. **How often did you encounter grief during calls?** (Multiple choice)
   - Almost never
   - Occasionally
   - Often
   - Almost all the time
6. **How often did you encounter anger during calls?** (Multiple choice)
   - Almost never
   - Occasionally
   - Often
   - Almost all the time
7. **May we use a quote from your responses about impact/mental health in the paper? Since this topic is sensitive, please also answer the following question.** (Multiple choice)
   - Yes
   - No
8. **If you have any specific instructions on what is and isn't ok to quote from your responses to this section, please indicate them here. If you would like for us to reach out to you about anything you mentioned, please also indicate that here and make sure to put your name in the first page of the survey.** (Long answer text)

Section 3: Weekly meetings

**SECTION 3**

1. **What did you think of the weekly meetings?** (Long answer text)
2. **May we use a quote from your responses about weekly meetings in the paper?** (Multiple choice)
   - Yes
   - No

Section 4: Messaging platform

**SECTION 4**

1. **What did you think of the messaging platform and how it was used?** (Long answer text)
2. **May we use a quote from your responses about the messaging platform in the paper?** (Multiple choice)
   - Yes
   - No

Section 5: Overall feedback

**SECTION 5**

1. **What advice would you like to share with future researchers who might conduct a similar study?** (Long answer text)
2. **Was there anything about how we conducted this study that worked well? (including from when you interviewed, trained, throughout when you were an interviewer in the study)** (Long answer text)
3. **Was there anything that did not work well? Why?** (Long answer text)
4. **Are there any other insights you would like to share?** (Long answer text)
5. **May we use a quote from your responses about overall feedback (this section) in the paper?** (Multiple choice)
   - Yes
   - No
